# Association of BCG vaccination policy and tuberculosis burden with incidence and mortality of COVID-19

**DOI:** 10.1101/2020.03.30.20048165

**Authors:** Giovanni Sala, Rik Chakraborti, Atsuhiko Ota, Tsuyoshi Miyakawa

**Affiliations:** Division of Systems Medical Science, Institute for Comprehensive Medical Science, Fujita Health University; Department of Economics, Christopher Newport University; Department of Public Health, Fujita Health University School of Medicine

## Abstract

**Background:** Evidence suggests non-specific benefits of the tuberculosis vaccine bacillus Calmette-Guérin (BCG) against non-related infections. Recent studies propose such protection may extend to the novel COVID-19 as well. This is a contested hypothesis.

**Methods:** Our ecological study confronts this hypothesis. We examine the effects of BCG vaccination on countries’ COVID-19 (a) cases and deaths (per million) and (b) exponential growth factors over specific periods of the pandemic. Since the BCG vaccine was derived from Mycobacterium bovis, a bacterium causing tuberculosis in cattle, having suffered from tuberculosis also may exert a non-specific protection against the COVID-19 as well. Along with BCG vaccination, we test the effect of the prevalence of tuberculosis.

We employ multiple regression and principal component analysis (PCA) to control for potentially confounding variables (*n* = 16).

**Results:** BCG vaccination policy and incidence of tuberculosis is associated with a reduction in both COVID-19 cases and deaths, and the effects of these two variables are additive (≈ 5% to 15% of total unique variance explained). The study of exponential growth factors in the initial stages of the pandemic further shows that BCG vaccination exerts a significant effect (up to 35% of unique variance explained).

**Conclusions:** Overall, these findings corroborate the hypothesis that BCG vaccination and exposure to tuberculosis may induce a non-specific protection against the novel SARS-CoV-2 infection, even after accounting for a large number of confounding influences. However, given the potential public-health benefits, our results indicate that the hypothesis deserves further attention and should not be hastily dismissed.

## Introduction

The recent outbreak of coronavirus disease 2019 (COVID-19) caused by the 2019 novel coronavirus (SARS-CoV-2) is undoubtedly among the gravest global threats the human race has confronted this millennium. But populations in different countries vary substantially in their experiences of risk of infection and mortality from this virus^1^. Take for instance, Japan, which, compared to many other countries, is physically and socially close to China, the source of the virus. Japan also belongs to the list of the countries that recorded the first case in the earliest phase of this pandemic. As of April 26^th^, while many governments have ‘locked-down’ numerous cities and entire countries in certain instances, Japanese authorities maintain relatively mild measures to mitigate the spread of the virus. They have simply advised citizens to stay home to the extent possible.

Nevertheless, most people in Japan, with the notable exception of the Tokyo metropolitan area, continue to work almost as usual. Surprisingly, despite the country’s head start in terms of infections (together with China and South Korea), the liberal measures taken, and the relatively low number of PCR tests conducted, as of May 10th, Japan ranks 116th and 96th among 199 countries and regions^1^, in cases and deaths, per million, respectively. Countries in Western and Eastern Europe also exhibit notable differences in COVID-19 prevalence and mortality. While many potential geographical, social, and biological differences including temperature, humidity, life expectancy, average income, social norms, and ethnic/genetic background could contribute to these observed variabilities among countries, these remain under examined. Successfully identifying such information could be a key step in the mitigation of the spread of the virus.

Studies report the close association of some vaccines (e.g., measles and Bacillus Calmette-Guérin (BCG)) with a lower risk of illness and death from other disorders^2–4^. Systematic reviews of epidemiological studies provide evidence for the non-specific beneficial effects of BCG vaccine on all-cause mortality^5,6^. Among these, of central interest is the induction of cytokines associated with trained immunity underlying such protection against non-related viral infections^7–9^.

Researchers in four countries will soon start a clinical trial of BCG vaccine on this disease, based on the proposed beneficial effect of BCG^9,4^. Popular interest in this topic spiked, interestingly, from the work of some commentators on the internet, including an esteemed blogger^10^, who first noticed a potential cross-country correlation between the administration of BCG vaccinations and the extent to which the spread of COVID-19 occurs. Subsequently, a number of ecological studies evaluated the hypothesis, with some finding results consistent^11–17^ and others finding results inconsistent^18–22^ with the idea.

In this study, we evaluate the hypothesis that BCG vaccination has protective effects against COVID-19 by utilizing publicly available datasets through multiple regression modeling. Our models consider factors such as a country’s mean temperature, percentage of population over 65 years of age, proportion of overweight people, and percentage of people living in urban areas. Finally, since BCG was originally derived as a bacterium from the one causing tuberculosis (TB), we also test the effects of TB prevalence, as represented by TB incidence in each country, on COVID-19 risk.

## Methods

### Selection of the countries

For all our analyses, we restricted our sample to countries with populations of at least one million for which at least 15 days of data since the detection of the first case were available as of April the 26^th^. These criteria ensure a sufficient number of data points to be reliable. We collected data for 142 countries which met these inclusion criteria. The details of our full dataset are reported in the supplemental materials section (Tables S1-3).

### Dependent variables

We examine three dependent variables: cases of COVID-19 per one million population (cases_pm_), deaths from COVID-19 per one million population (deaths_pm_), and growth factors of COVID-19-related cases and deaths.

The above results referred to the total cases and deaths per one million population at a specific date (as of April the 26^th^). Instead of using a fixed date for all the countries regardless on when the epidemic started, the present analysis employed a fixed number of days from the day a specific threshold of cases was reached. We thus examined the growth factors for each country’s COVID-19 cases/deaths.

The relationship between time and cumulative number of cases follows an exponential function in the initial phases of an epidemic^23^.

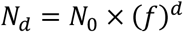

where *N_d_* and *N*_0_ are the cumulative number of cases/deaths at day *d* and day 0, respectively, and *f* is the growth factor. This analysis aimed to assess the impact of BCG policy and TB incidents on the growth factor.

The relationship was linearized by applying a logarithmic transformation.

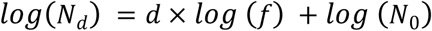

Then, a simple linear regression was run to extract the coefficient *log (f*), which was used as the dependent variable in the analyses:

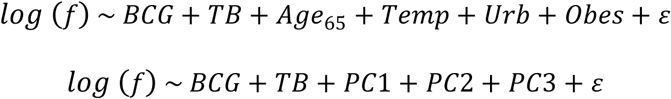

as in the previous models.

Since selecting a specific criterion for the extraction of the growth factors is to some extent arbitrary, we ran two distinct sets of analyses with different inclusion criteria. First, we selected the data from the first 15 days after reaching *1 case and 1 death per one million population* in each country. Second, we ran a sensitivity analysis by selecting the data from the first 15 days after reaching the *100^th^ absolute case and the 30^th^ death* in each country.

Both the analyses included a vector of weights (weighted linear regression). The weights were the *R^2^* statistics calculated in the single regression for the estimation of *log (f)*. This approach allowed us to reduce the weight of those countries that did not show a nearly perfect exponential relationship between recorded cases/deaths and days in the regression analysis.

### Independent variables of interest

We investigate the relationships between the dependent variables and (a) the country’s BCG vaccination policy and (b) the mean incidence of tuberculosis per 100,000 people between 2010 and 2018 (hereafter, TB incidence)^24^.

Furthermore, we evaluate the potential impact of the type of BCG strains on the COVID-19 cases_pm_ and deaths_pm_. In accordance with Zhang et al.^25^, we classified the BCG strains into two categories: G3+, those strains that lost 28 “Group 3” epitopes (Denmark, Pasteur and others); and G3−, those strains that still contain these epitopes (Tokyo, Russia and Moreau). We could retrieve this information for 69 countries^26–28^ (See Table S9 for details). This variable was confounded with the country’s BCG vaccination policy (*χ^2^* = 10.659, *p* = .0011; i.e. The countries that discontinued mandatory BCG vaccination tend to use G3+ strains; Table S1). Thus, we investigated the effects of the type of BCG strains in a separate analysis.

### Control variables

The observed effect of a country’s BCG vaccination policy and TB incidence may be an artifact due to some confounding variables. First, we chose four covariates, listed below, as controls to test whether the impact of the independent variables of interest remained significant accounting for these potentially confounding influences. We chose 19 potentially confounding factors and created a correlation matrix (Table S1). For the rational to choose them, see Supplementary Discussion for details. Many of these variables are correlated with each other and, to avoid collinearity-related issues, we selected following four variables, that are suspected to have relatively a direct causal effect on COVID-19 risk for the following reasons.

- The country’s proportion of individuals over 65 years of age: Older age is a risk factor for mortality of patients with COVID-19^29,30^. People who are 65 or older constitute approximately 80% share of US COVID-19 deaths, as of May 6^th 31^; People who are 60 or older constitute more than 93 % share of COVID-19 deaths in Japan^32^. We expect higher percentage of population over 65 years of age would increase the number of cases and deaths per population. The countries that discontinued or never had mandatory BCG vaccination policy (BCG N countries) tend to be developed countries whose percentage of elderly people might be relatively high (Figure 3a, b) and this could be a major confounding factor^19^.
- The country’s mean temperature in February and March 2020: Like other coronaviruses and respiratory infections, the spread of COVID-19 may be hindered by higher temperature and ecological studies have reported negative correlations between temperature and COVID-19 indices^17,33,34^. Since BCG N countries tend to have lower average temperature compared to BCG Y countries during this pandemic (Figure S13a, c), temperature could be a major confounding factor.
- The country’s percentage of population living in urban centers: The virus is more likely to spread among crowded urban centers than countryside locations. Since BCG N countries are mostly in Western Europe, whose percentage of urban population tend to be high (Table S1; Figure 4a, b), this could be a potentially serious confounding factor.
- The country’s percentage of population that suffers from obesity (BMI > 25): In fact, obesity is a well-known general risk factor for a number of clinical conditions. Obesity and its related diseases are considered to be a risk factor of COVID-19^35–37^. BCG N countries tend to have high percentage of obese population^19^ (Table S1; Figure S6). Thus, being overweight may be associated, for instance, with a higher death rate from COVID-19 infections.

We were able to retrieve the complete set of data regarding these variables from a total of 117 countries.

Fifteen additional control variables were used for correlation and subpopulation analyses.

Twelve of them had sufficient data to be evaluated for their possible confounding effects in the PCA and multiple regression analyses. Table 1 presents a summary of these variables. See Supplementary Discussion for the details about these variables.

**Table 1.**
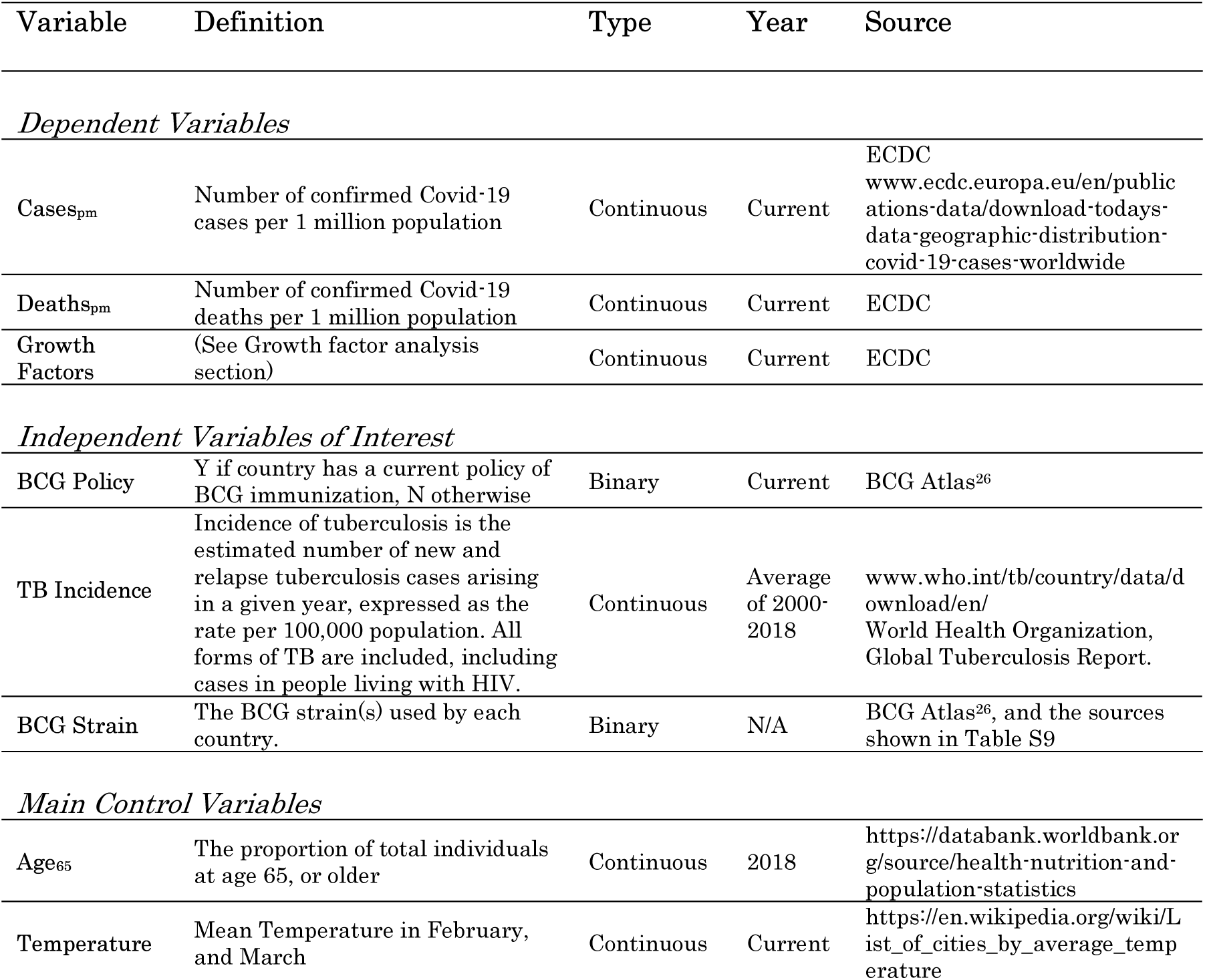
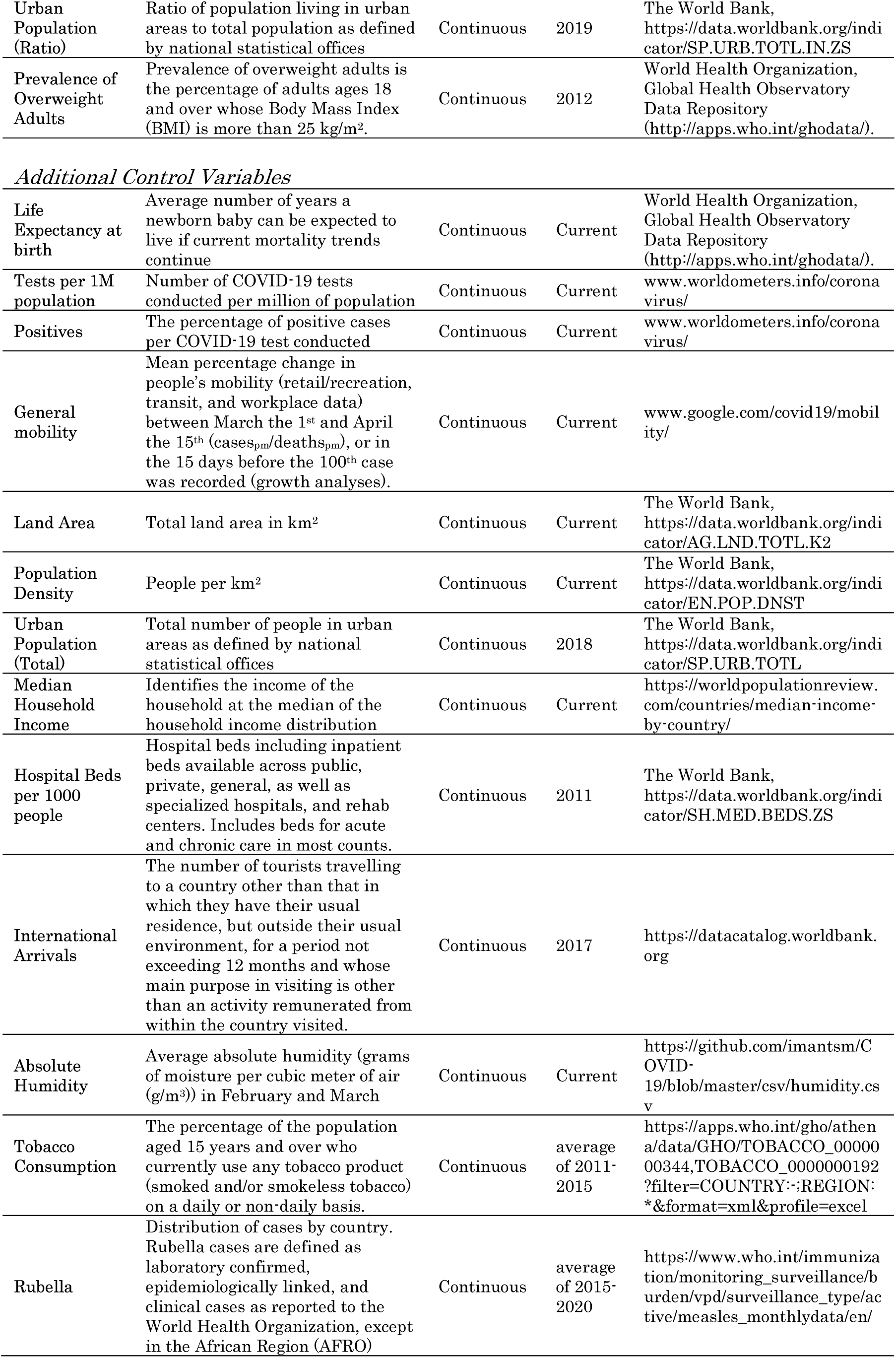
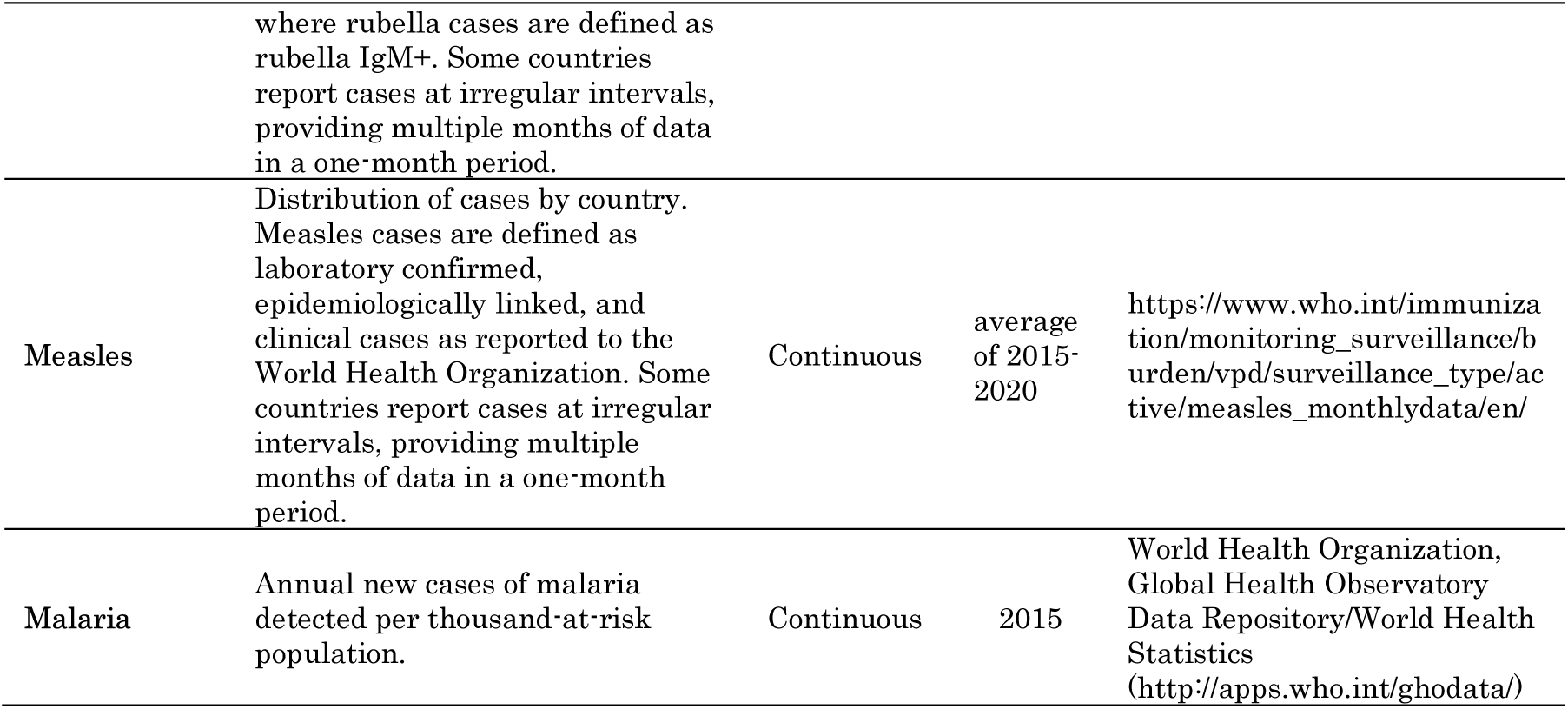
Description of the variables employed in the analyses.

| Variable | Definition | Type | Year | Source |
| --- | --- | --- | --- | --- |
| *Dependent Variables* |  |  |  |  |
| **Cases<sub>pm</sub>** | Number of confirmed Covid-19 cases per 1 million population | Continuous | Current | ECDC<br>www.ecdc.europa.eu/en/publications-data/download-todays-data-geographic-distribution-covid-19-cases-worldwide |
| **Deaths<sub>pm</sub>** | Number of confirmed Covid-19 deaths per 1 million population | Continuous | Current | ECDC |
| **Growth Factors** | (See Growth factor analysis section) | Continuous | Current | ECDC |
| *Independent Variables of Interest* |  |  |  |  |
| **BCG Policy** | Y if country has a current policy of BCG immunization, N otherwise | Binary | Current | BCG Atlas<sup>26</sup> |
| **TB Incidence** | Incidence of tuberculosis is the estimated number of new and relapse tuberculosis cases arising in a given year, expressed as the rate per 100,000 population. All forms of TB are included, including cases in people living with HIV. | Continuous | Average of 2000-2018 | www.who.int/tb/country/data/download/en/ World Health Organization, Global Tuberculosis Report. |
| **BCG Strain** | The BCG strain(s) used by each country. | Binary | N/A | BCG Atlas<sup>26</sup>, and the sources shown in Table S9 |
| *Main Control Variables* |  |  |  |  |
| **Age<sub>65</sub>** | The proportion of total individuals at age 65, or older | Continuous | 2018 | https://databank.worldbank.org/source/health-nutrition-and-population-statistics |
| **Temperature** | Mean Temperature in February, and March | Continuous | Current | https://en.wikipedia.org/wiki/List_of_cities_by_average_temperature |
| **Urban Population (Ratio)** | Ratio of population living in urban areas to total population as defined by national statistical offices | Continuous | 2019 | The World Bank, https://data.worldbank.org/indicator/SP.URB.TOTL.IN.ZS |
| **Prevalence of Overweight Adults** | Prevalence of overweight adults is the percentage of adults ages 18 and over whose Body Mass Index (BMI) is more than 25 kg/m<sup>2</sup>. | Continuous | 2012 | World Health Organization, Global Health Observatory Data Repository (http://apps.who.int/ghodata/). |
| *Additional Control Variables* |  |  |  |  |
| **Life Expectancy at birth** | Average number of years a newborn baby can be expected to live if current mortality trends continue | Continuous | Current | World Health Organization, Global Health Observatory Data Repository (http://apps.who.int/ghodata/). |
| **Tests per 1M population** | Number of COVID-19 tests conducted per million of population | Continuous | Current | www.worldometers.info/coronavirus/ |
| **Positives** | The percentage of positive cases per COVID-19 test conducted | Continuous | Current | www.worldometers.info/coronavirus/ |
| **General mobility** | Mean percentage change in people's mobility (retail/recreation, transit, and workplace data) between March the 1<sup>st</sup> and April the 15<sup>th</sup> (cases<sub>pm</sub>/deaths<sub>pm</sub>), or in the 15 days before the 100<sup>th</sup> case was recorded (growth analyses). | Continuous | Current | www.google.com/covid19/mobility/ |
| **Land Area** | Total land area in km<sup>2</sup> | Continuous | Current | The World Bank, https://data.worldbank.org/indicator/AG.LND.TOTL.K2 |
| **Population Density** | People per km<sup>2</sup> | Continuous | Current | The World Bank, https://data.worldbank.org/indicator/EN.POP.DNST |
| **Urban Population (Total)** | Total number of people in urban areas as defined by national statistical offices | Continuous | 2018 | The World Bank, https://data.worldbank.org/indicator/SP.URB.TOTL |
| **Median Household Income** | Identifies the income of the household at the median of the household income distribution | Continuous | Current | https://worldpopulationreview.com/countries/median-income-by-country/ |
| **Hospital Beds per 1000 people** | Hospital beds including inpatient beds available across public, private, general, as well as specialized hospitals, and rehab centers. Includes beds for acute and chronic care in most counts. | Continuous | 2011 | The World Bank, https://data.worldbank.org/indicator/SH.MED.BEDS.ZS |
| **International Arrivals** | The number of tourists travelling to a country other than that in which they have their usual residence, but outside their usual environment, for a period not exceeding 12 months and whose main purpose in visiting is other than an activity remunerated from within the country visited. | Continuous | 2017 | https://datacatalog.worldbank.org |
| **Absolute Humidity** | Average absolute humidity (grams of moisture per cubic meter of air (g/m<sup>3</sup>)) in February and March | Continuous | Current | https://github.com/imantsm/COVID-19/blob/master/csv/humidity.csv |
| **Tobacco Consumption** | The percentage of the population aged 15 years and over who currently use any tobacco product (smoked and/or smokeless tobacco) on a daily or non-daily basis. | Continuous | average of 2011-2015 | https://apps.who.int/gho/athena/data/GHO/TOBACCO_000000344.TOBACCO_0000000192?filter=COUNTRY:-:REGION:*&format=xml&profile=excel |
| **Rubella** | Distribution of cases by country. Rubella cases are defined as laboratory confirmed, epidemiologically linked, and clinical cases as reported to the World Health Organization, except in the African Region (AFRO) | Continuous | average of 2015-2020 | https://www.who.int/immunization/monitoring_surveillance/burden/vpd/surveillance_type/active/measles_monthlydata/en/ |
|  | where rubella cases are defined as rubella IgM+. Some countries report cases at irregular intervals, providing multiple months of data in a one-month period. |  |  |  |
| **Measles** | Distribution of cases by country. Measles cases are defined as laboratory confirmed, epidemiologically linked, and clinical cases as reported to the World Health Organization. Some countries report cases at irregular intervals, providing multiple months of data in a one-month period. | Continuous | average of 2015-2020 | https://www.who.int/immunization/monitoring_surveillance/burden/vpd/surveillance_type/active/measles_monthlydata/en/ |
| **Malaria** | Annual new cases of malaria detected per thousand-at-risk population. | Continuous | 2015 | World Health Organization, Global Health Observatory Data Repository/World Health Statistics (http://apps.who.int/ghodata/) |

### Preprocessing of the variables

The continuous variable of interest (TB incidence) was scaled (i.e., z-scores were calculated). Also, the Ordered Quantile (ORQ) normalization transformation^38^ was used to normalize and standardize the other continuous variables (e.g., cases_pm_, deaths_pm_, and the control variables) which did not show a normal distribution. This transformation is necessary to satisfy a fundamental statistical assumption of linear modeling: normally distributed residuals. This procedure was also useful to mitigate potentially confounding effects of outliers, especially in highly skewed and dispersed data.

### Analytic strategy

First, we examined the correlation matrix (Table S2) as a preliminary exploration of the relationships of interest. We explored scatter plots for COVID-19 cases_pm_/deaths_pm_ by each of our 16 control variables (see Supplementary Figures S1 - S17). For each potential confounder, we selected a subset of countries with and without universal BCG policies so that these countries were essentially identical by mean values for the confounding variable. This approach permits a visual comparison of COVID-19 incidence and mortality across countries that implement and do not implement a universal BCG vaccination policy but are similar in terms of the potential confounder. These comparisons were depicted in boxplots (Supplementary Figures)

The total number of observations (countries) limited the inclusion of all confounding variables in a single multiple regression analysis (high dimensionality). Further, these control variables were sometimes significantly correlated to each other (i.e., multicollinearity). In order to resolve both these problems, we ran (a) a series of multiple regression analyses including the main control variables and (b) a Principal Component Analysis (PCA) to extract a few components from all the control variables.

We ran two multiple regression analyses to evaluate the effects of the IVs of interest (BCG policy and TB incidence) on the number of COVID-19 cases and deaths per one million population (as of April the 26th). We included four main control variables as covariates:

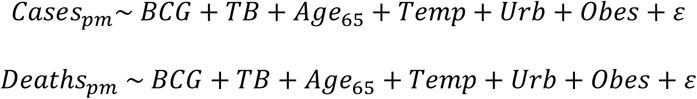

where *BCG* and *TB* represent a country’s vaccination policy and TB incidence, *Age_65_* represents the proportion of population over 65 years of age, *Temp* represents the mean temperature in February and March 2020, *Urb* represents the proportion of the population living in urban centers, *Obes* represents the percentage of population that suffers from obesity (BMI > 25), and *ε* captures the model’s residual error.

We also run a regression subset selection^39^ to identify the optimal set of variables based on the Bayesian Information Criterion (BIC). After applying this algorithm, we calculated the variables’ standardized beta (*β*) coefficients and percentages of unique explained variances^40,41^ for each independent variable (predictors). Beta coefficients capture standardized impacts: they measure the effect of a one- standard-deviation change in each predictor on the dependent variable. We report these coefficients to present the relative impacts in a common comparable metric for each predictor. The unique explained variance is the portion of the dependent variable’s variation that can be accounted solely by the variation of each predictor. These estimates summarize the unique impact of each predictor after controlling for the impacts of all other predictors included in the regression.

As a final step, we employ PCA as a stress-test for the results of the multiple regression analysis. We applied an orthogonal extraction (varimax) to the 12 control variables and estimated three principal components (estimated by k-fold cross validation) that explained about 70-75% of the total variance. The three components (PC1, PC2, and PC3) replaced the four main control variables in the multiple regression models.

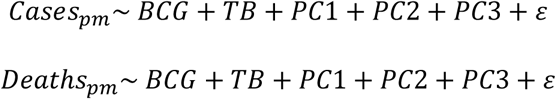

Since, we were able to retrieve the complete set of data for small number of countries (*N=* 53), we used an imputation algorithm prior to running the PCA.^42,43^

## Results

### Correlations between variables and subpopulation analyses

Table S4 presents the correlation matrix between the number of cases_pm_ and deaths_pm_ and the control variables is shown in. Thirteen out of 16 variables evaluated showed high or moderate correlations with COVID-19 cases_pm_/ deaths_pm_. For each potentially confounding variable, we selected a subset of the observations (countries) so that its mean value was approximately the same for the subset of selected countries which includes both implementers (BCG = Y) and non-implementers (BCG = N) of a universal BCG vaccination policy. Legends of Figure 2, 3, 4 and S1-S17 describe the selection criteria. Table S6 shows means ± SDs for each variable and BCG group after the selection.

**Figure 1.**
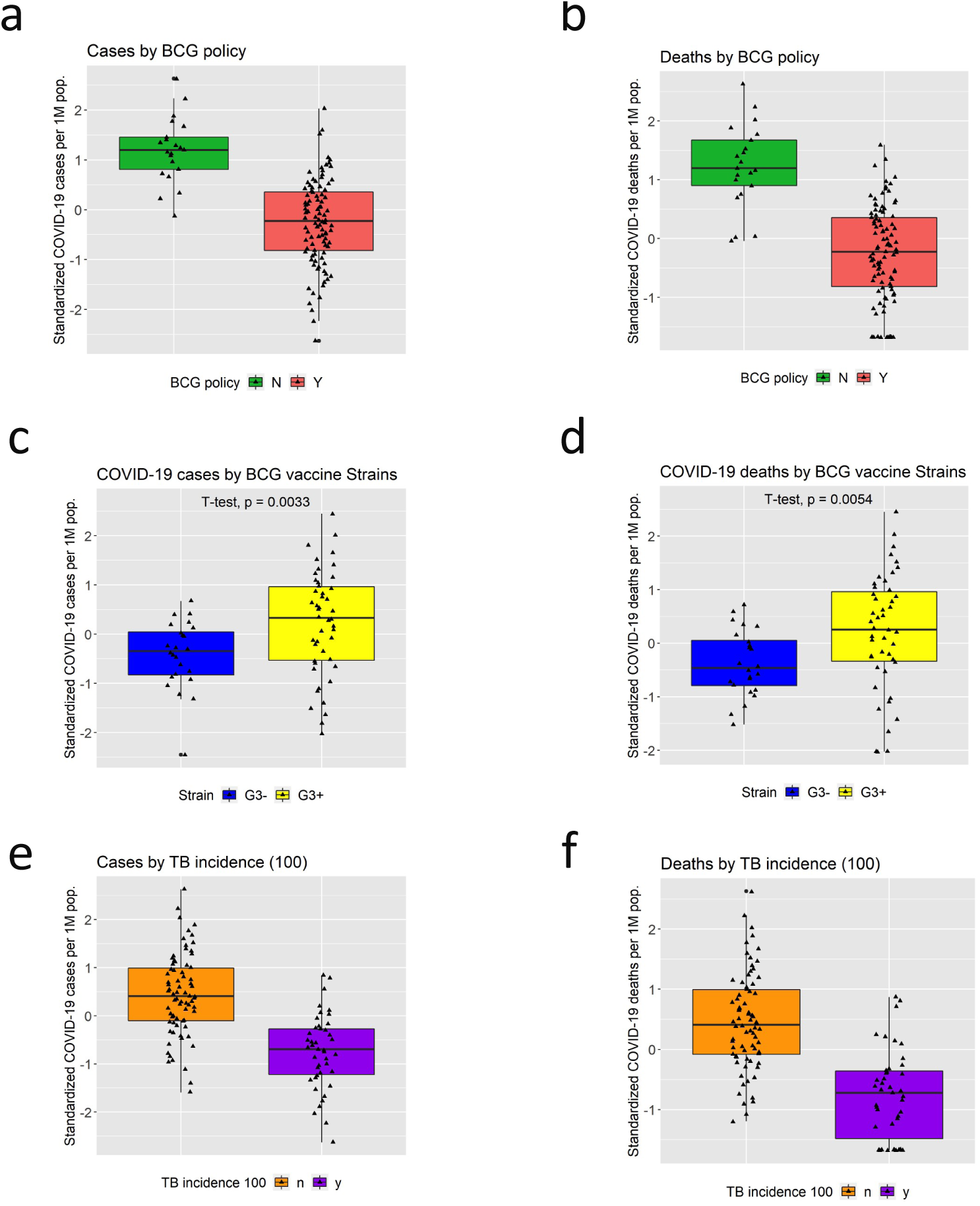
Effect of BCG vaccination policy, BCG strain used, and TB burden on COVID-19 a, b. The boxplot of total cases (a) and deaths (b) per one million population sorted by BCG Group in countries (N= 117). Group N (no current BCG vaccination) show a significantly higher rate of cases and deaths of COVID-19 compared to Group Y (countries currently implementing BCG vaccination). c, d. The boxplot of total cases (c) and deaths (d) per one million population sorted by BCG strains used in countries (N= 69). G3+ group (Denmark, Pasteur, Connaught, China, Tice and the one by Intervax Ltd Toronto) show a significantly higher rate of cases and deaths of COVID-19 compared to G3− group (Tokyo, Russia/Bulgaria, and Moreau). e, f. The boxplot of total cases (c) and deaths (d) per one million population sorted by TB burden in countries (N= 117). Low TB burden countries group show a significantly higher rate of cases and deaths of COVID-19 compared to high TB burden countries. The vertical lines of box plots are the range of values that are not outliers (the definition of outliers should be Q3/Q1 ±1.5 × interquartile range). The horizontal lines are the median, and the two quartiles (Q1 and Q3).

**Figure 2.**
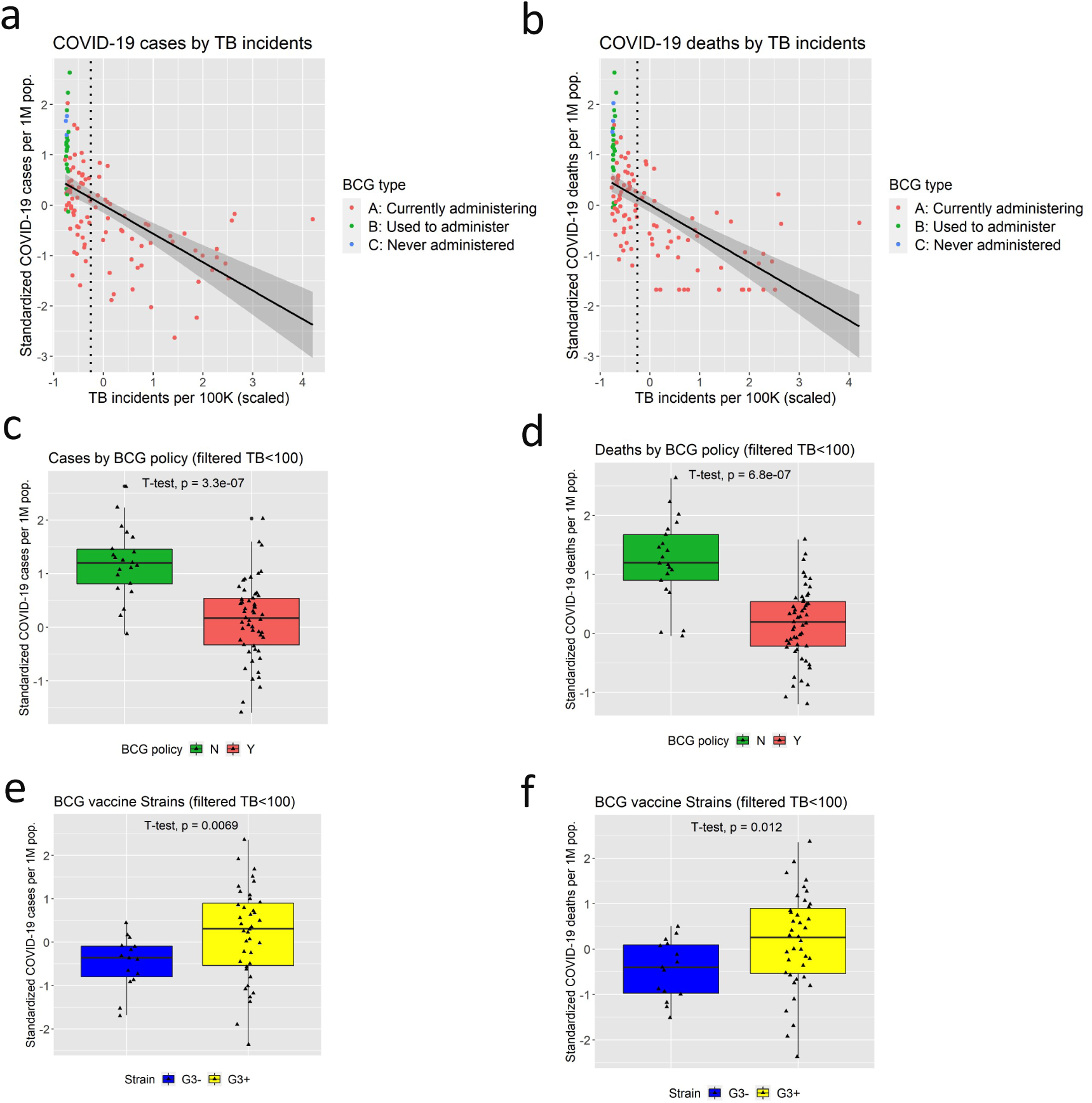
TB incidence and COVID-19: Scatter plot of total cases (a) or deaths (b) per one million population by TB incidents (per 10K population; average from 2000 to 2018). The vertical dotted line indicates the threshold of 78 years, that is subsequently used for the analyses shown in the panel c-f. The boxplot of total cases (c) or deaths (d) per one million population sorted by BCG Group in countries with life expectancy higher than 78 years. Group N show a significantly higher rate of cases or deaths of COVID-19 compared to Group Y. The boxplot of total cases (e) or deaths (f) per one million population sorted by BCG strain types in countries with life expectancy higher than 78 years. G3+ group show a significantly higher rate of or and deaths of COVID-19 compared to Group G3−.

**Figure 3.**
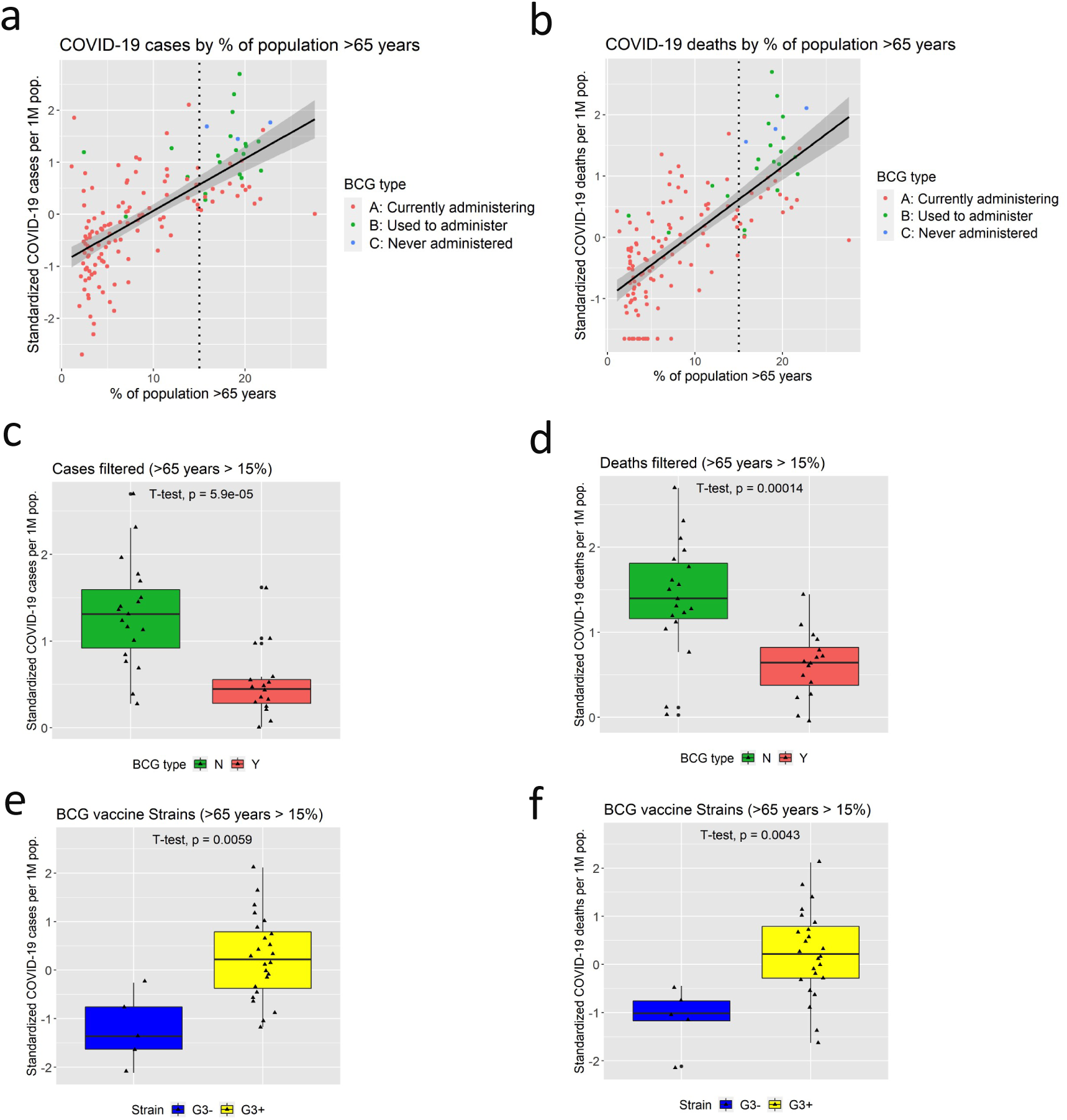
Percentage of population aged 65 and above and COVID-19: Scatter plot of total cases (a) or deaths (b) per one million population by Percentage of population aged 65. The vertical dotted line indicates the threshold of 15 %, that is subsequently used for the analyses shown in the panel c-f. The boxplot of total cases (c) or deaths (d) per one million population sorted by BCG Group in countries with percentage of population aged 65 and above higher than 15%. Group N show a significantly higher rate of cases or deaths of COVID-19 compared to Group Y. The boxplot of total cases (e) or deaths (f) per one million population sorted by BCG strain types in countries with percentage of population aged 65 and above higher than 15%. G3+ group show a significantly higher rate of cases or deaths of COVID-19 compared to Group G3−.

**Figure 4.**
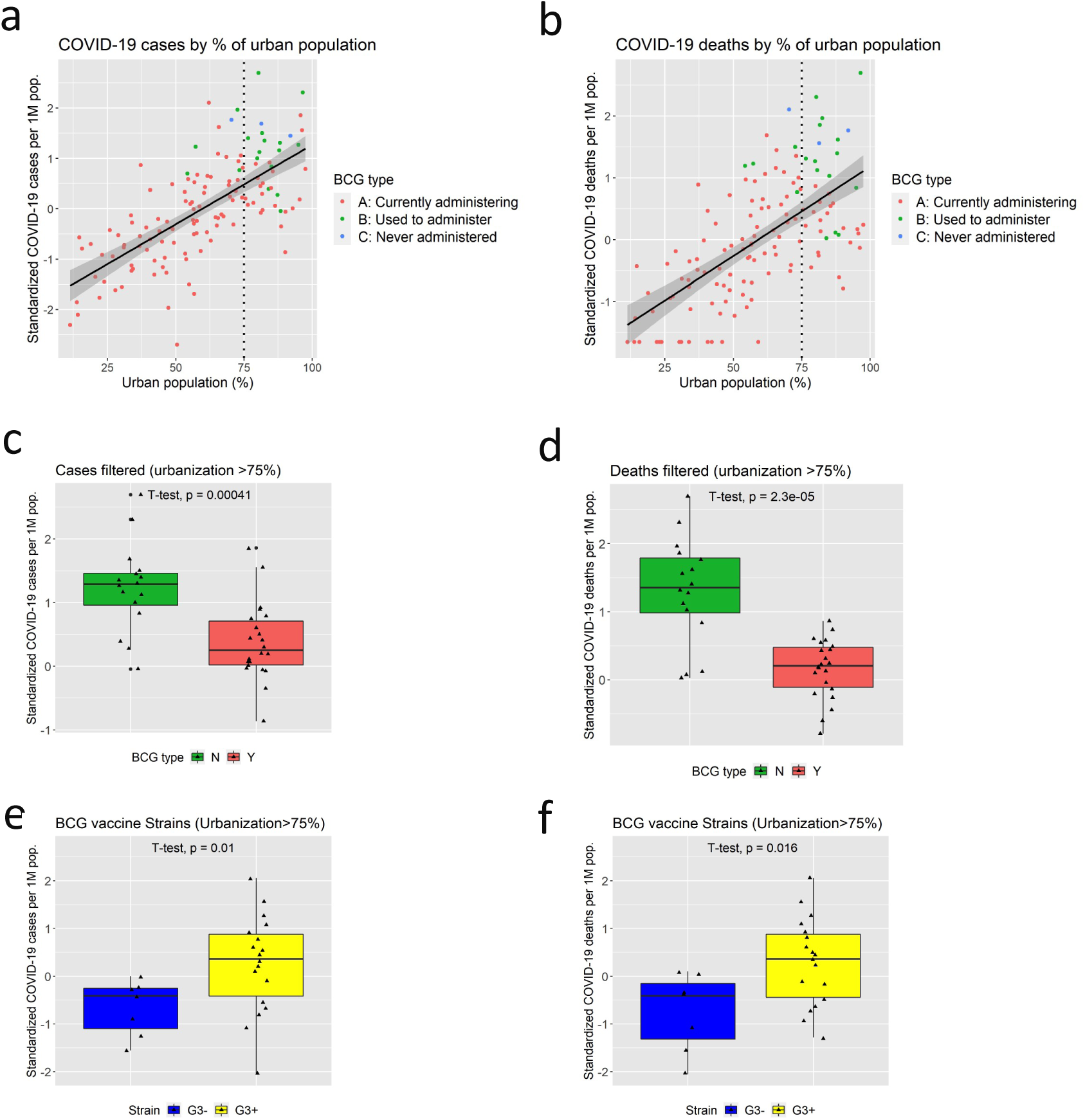
Percentage of urban population as a fraction of total population: Scatter plot of total cases (a) or deaths (b) per one million population by urban population as a fraction of total population (%). The vertical dotted line indicates the threshold of 75 %, that is subsequently used for the analyses shown in the panel c-f. The boxplot of total cases (c) or deaths (d) per one million population sorted by BCG Group in countries with fraction of urban population higher than 75%. Group N show a significantly higher rate of cases and deaths of COVID-19 compared to Group Y. The boxplot of total cases (e) or deaths (f) per one million population sorted by BCG strain types in countries with fraction of urban population higher than 75%. G3+ group show a significantly higher rate of cases or deaths of COVID-19 compared to Group G3−.

Figures 2, 3, 4 and S1-S17 provide scatterplots showing the observed association between COVID-19 cases_pm_/deaths_pm_ by TB incidence, the 16 control variables and 3 additional variables. Figure 2, 3, 4 and Figures S1-S17 present box plots depicting comparisons between BCG groups. Among the subset of the countries in each comparison with similar mean values of potential confounding variables, these figures indicate that numbers of COVID-19 cases_pm_/ deaths_pm_ for countries with a BCG vaccination policy are significantly smaller (See details in Figure 2, 3, 4 and Figures S1-S17 and their legends). The COVID-19 cases_pm_ for median household income (Figure S2b; *p* = 0.073) is the sole exception. These results suggest that each potential confounding variable did not solely account for the observed effect of the BCG vaccination on the COVID-19 risk.

### Cases and Deaths per one million population

Regarding total cases_pm_, the linear model showed a statistically significant effect of the country’s BCG vaccination policy (*p* = .0026) and a marginally statistically significant effect of TB incidence (*p* = .0504). These results indicated that the countries that implemented a universal BCG vaccination policy and exhibited more TB incidents are associated with fewer COVID-19 cases per one million population. Population ages 65 and above (% of total population; Age_65_) and urban population (% of total population) were significant as well (*p* = .0045 and, *p* = .0001, respectively). Temperature and obesity did not exert a significant effect (*p* = .9393 and *p* = .1978, respectively). The model’s adjusted *R^2^* was .6202 (i.e., approximately 62% of the observed variance was explained). The inspection of the model’s residuals did not reveal any anomaly (Shapiro-Wilk test *p* = .2948). See also Figures 1a, 1e, 2a, 2c, 3a, 3c, 4a, and 4c. The boxplots and scatterplots depict the relationship between the statistically significant independent variables and cases_pm_/deaths_pm_ (in both the whole sample and subgroups). A very similar pattern of results was obtained with deaths_pm_. The model’s residuals were again normally distributed (*p*= .8919). See also Figures 1b, 1f, 2b, 2d, 3b, 3d, 4b, and 4d (as above).

Including a large number of variables in linear multiple regression analysis can be problematic when the number of observations is limited, as is the case here. To respond to this problem, we used a regression subset selection algorithm to identify the optimal set of variables according to the BIC criterion. Consistent with the above analysis, four significant variables were selected. We calculated the Beta coefficient (*β*) estimates to measure the magnitude of the effect of each variable. Finally, we estimated the amount of unique variance – i.e., the variance explained by a certain variable by controlling for all the other variables in the model – accounted for by the included variables. Table 2 summarizes the results.

**Table 2.** The linear model of cases and deaths per one million population.

| Variable | $\beta$ Coefficient | $p$-value | Unique Variance |
| --- | --- | --- | --- |
| *Cases<sub>pm</sub>* |  |  |  |
| *BCG Policy* | -0.2180 | .0016 | 7.70% |
| *TB Incidence* | -0.1593 | .0279 | 3.39% |
| *Age<sub>65</sub>* | 0.2839 | .0002 | 11.17% |
| *Urbanization (%)* | 0.3599 | $2.03 \times 10^{-6}$ | 17.59% |
| ***Deaths<sub>pm</sub>*** |  |  |  |
| *BCG Policy* | −0.2478 | .0003 | 10.43% |
| *TB Incidence* | −0.1992 | .0051 | 5.95% |
| *Age<sub>65</sub>* | 0.3358 | $6.67 \times 10^{-6}$ | 15.89% |
| *Urbanization (%)* | 0.2587 | .0003 | 10.06% |

Finally, a Principal Component Analysis (PCA) was run to stress-test the above results. Table S8 shows the component loadings (*λ*s) for each variable included in each of the three analyses (the present one and two growth analyses). The three components were then added to a multiple regression model along with the two independent variables of interest. For cases_pm_, the country’s BCG policy was statistically significant (*p* = .0173), while TB incidence was marginally significant (*p* = .0773). We find a similar pattern of results for deaths_pm_. In this case, however, both the country’s BCG policy and TB incidence are statistically significant (*p* = .0017 and *p* = .0023). Table 3 summarizes these results.

**Table 3.** The linear model of cases and deaths per one million population with the PCA components.

| Variable | β-Coefficient | $p$-value | Unique Variance |
| --- | --- | --- | --- |
| ***Cases<sub>pm</sub>*** |  |  |  |
| *BCG Policy* | −0.1449 | .0173 | 3.39% |
| *TB Incidence* | −0.1095 | .0773 | 1.56% |
| *PC1* | 0.6300 | $9.46 \times 10^{-16}$ | 37.43% |
| *PC2* | 0.1401 | .0065 | 4.63% |
| *PC3* | 0.1130 | .0274 | 2.82% |
| ***Deaths<sub>pm</sub>*** |  |  |  |
| *BCG Policy* | −0.1920 | .0017 | 6.31% |
| *TB Incidence* | −0.1905 | .0023 | 5.92% |
| *PC1* | 0.5207 | $6.95 \times 10^{-12}$ | 28.95% |
| *PC2* | 0.0812 | .1107 | 1.14% |
| *PC3* | 0.2172 | $3.34 \times 10^{-5}$ | 11.28 % |

Incidentally, it is worth noting that some PCA components are extracted from many variables (*n* = 12). Thus, the components are more likely to be collinear with the independent variables of interest. This may explain the slight reduction in the effect size (*β*) of BCG Policy and TB effect size compared to the results reported in Table 2. For more details, see the R codes in the supplemental materials.

### Strain Analysis

Along with the effects of the BCG policy and the rate of TB incidents, we examined whether the strains of the vaccine used exerted a further impact. Specifically, the G3+ strains of the BCG vaccine are the mildest among the BCG strains (i.e., contain most deletion in their genome). These strains were therefore supposed to provide the least non-specific protective effect against COVID-19 infection (both in terms of cases and deaths).

We compared the two types of strains with a Welch’s *t*-test. The G3+ strains were associated to a higher number of both cases_pm_ and deaths_pm_ (*t* = −3.047, *df=* 64.647, *p*-value = 0.0033 and *t* = −2.872, *df=* 67.530, *p*-value = 0.0054, respectively). See also Figures 1c-d, 2e-f, 3e-f, and 4e-f. These boxplots depict the differences between the strains in cases_pm_/deaths_pm_ (both the whole sample and subgroups).

### Growth factor analysis

In this set of multiple regression models − the ones with the thresholds of 1 case and 1 death per one million population − the median of *R^2^* values was .9604 and .9544 for cases and deaths. This result indicated that (a) the assumption of the exponential relationship between days and cumulative cases_pm_/deaths_pm_ was substantially met and (b) the selection criterion for the thresholds was satisfactory. The number of complete observations for cases and deaths were N= 109 and N= 66, respectively. The country’s BCG policy was a significant predictor of the growth factor of both the COVID-19 cases and deaths (*p* = .0038 and *p* = .0069). By contrast, TB incidence was not significant (*p*s ≥ .7542). Age65 was significant (*p*s ≤ .0083). All the other variables (urbanization, temperature, and obesity) were nonsignificant (*p*s ≥ .5386). Finally, the model’s residuals followed a normal distribution (*p* ≥ .9609). See also Figures 5a and 5b that depict the average slope of the COVID-19 cases and deaths throughout the 15-day period in BCG in implementers (BCG = Y) and non-implementers (BCG = N).

**Figure 5.**
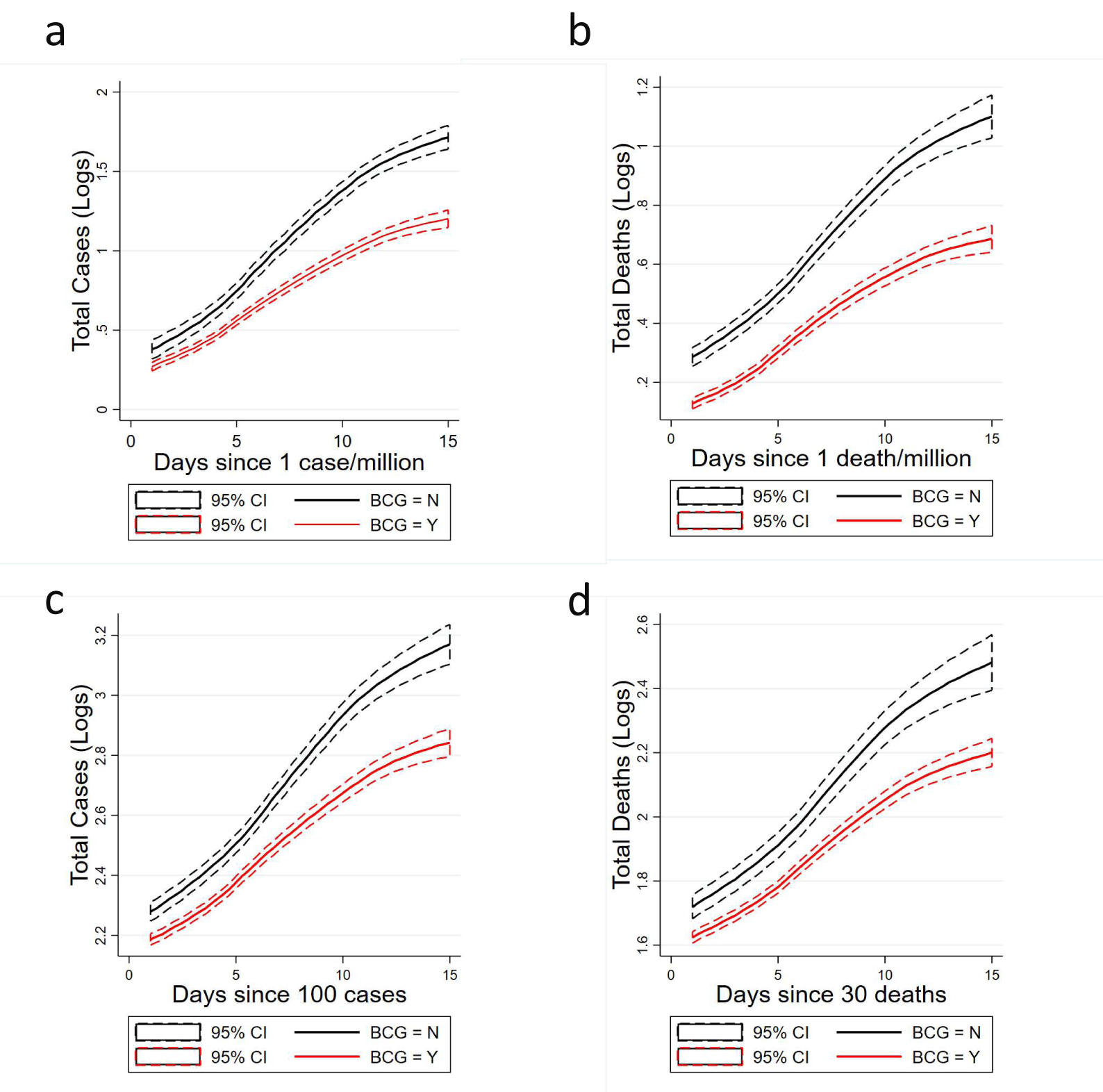
Growth curves: mean log-transformed total cases and deaths per one million population by BCG Group as a function of day in the first growth factor analysis (a and b) and in the second growth factor analysis (c and d). Group N show a significantly higher growth rate of both cases and deaths of COVID-19 compared to Group Y. Note that the starting points (Day 1) of the curves are slightly different. This discrepancy may simply represent that the overall trend occurs earlier then Day 1 and does not have meaningful impact on the growth factors. For instance, assume that a BCG Y country at Day 0 reports 95 cumulative cases, and at Day 1 reports 105 (+10), whereas a country BCG N at Day 0 reports 95 cases but at Day 1, since its growth factor is bigger, may reports 120 cases (+25). Hence, the difference at Day 1.

The regression subset selection model retained the two significant variables. The results are summarized in Table 4.

**Table 4.**
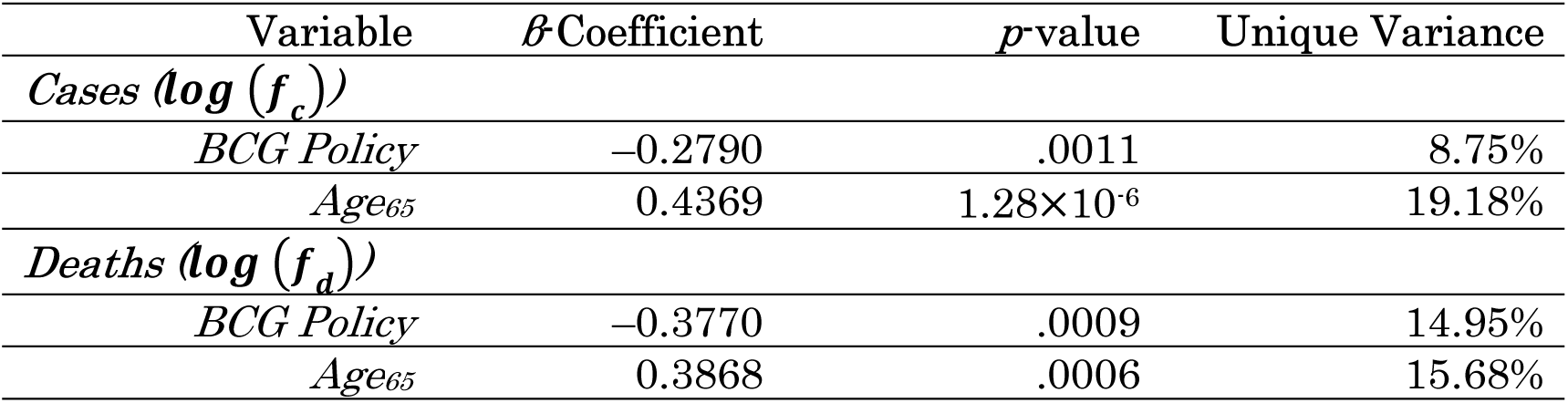
Models of growth factor in cases and deaths per one million population.

Finally, the results of PCA (*N*= 131 and *N*= 76 for cases and deaths, respectively, after missing data imputation) showed the same tendency. BCG policy was a predictor of the growth factor in both the country’s recorded cases and deaths (*p* = .0223 and *p* = .0492, respectively). TB incidence was not significant (*p*s ≥ .4136). The inspection of the models’ residuals did not show any issues (*p*s ≥ .1068). The results are summarized in Table 5.

**Table 5.** Models of growth factor in cases and deaths per one million population with the PCA components.

| Variable | $\beta$ Coefficient | $p$ value | Unique Variance |
| --- | --- | --- | --- |
| ***Cases<sub>pm</sub>*** |  |  |  |
| *BCG Policy* | −0.1906 | .0263 | 3.12% |
| *TB Incidence* | −0.0725 | .4136 | 0% |
| *PC1* | 0.4119 | 4.01×10<sup>−5</sup> | 11.97% |
| *PC2* | 0.0456 | .5249 | 0% |
| *PC3* | 0.0231 | .7449 | 0% |
| ***Deaths<sub>pm</sub>*** |  |  |  |
| *BCG Policy* | −0.2419 | .0492 | 4.06% |
| *TB Incidence* | −0.0851 | .4209 | 0% |
| *PC1* | 0.3632 | .0051 | 9.38% |
| *PC2* | 0.1067 | .2671 | 0.35% |
| *PC3* | 0.1417 | .1497 | 1.55% |

Then, we analyzed the growth factors of those countries that recorded at least 15 days of data after the 100^th^ absolute COVID-19 case and/or the 30^th^ death. Since the absolute cases/deaths were used, this analysis also included the country’s total population as a covariate. Again, the models’ residuals followed a normal distribution (*p*s ≥ .3592). The median of *R^2^* values was .9714 and .9776 for the cases and deaths, respectively. As mentioned above, this outcome indicated that the assumption of the exponential relationship between days and cumulative cases/death was substantially met and that the chosen thresholds performed satisfactory. The number of complete observations for cases and deaths were *N=* 94 and *N*= 52, respectively. The country’s BCG policy (*p* = .0040 and *p* =.0017) and total population (*p* = 2.15×10^−8^ and *p* = 1.26×10^−5^) were the only two significant predictors.

The regression subset selection model retained three variables for the growth factors of the cases, one of which was the country’s BCG vaccination policy. Temperature was significant as well. Only two variables were retained in the models of deaths. See also Figures 5c and 5d (as above). The results are summarized in Table 6.

**Table 6.** Models of growth factor in cumulative absolute cases and deaths.

| Variable | $\beta$ Coefficient | $p$ value | Unique Variance |
| --- | --- | --- | --- |
| ***Cases (log($f_c$))*** |  |  |  |
| *BCG Policy* | −0.3689 | 3.25×10<sup>−5</sup> | 16.63% |
| *Total Population* | 0.4605 | 1.69×10<sup>−7</sup> | 25.52% |
| *Temperature* | 0.3246 | .0004 | 12.22% |
| ***Deaths (log($f_d$))*** |  |  |  |
| *BCG Policy* | −0.5800 | 2.85×10<sup>−6</sup> | 35.05% |
| *Total Population* | 0.5161 | 2.09×10<sup>−5</sup> | 29.74% |

The mean growth factor for the cases (*f_c_*) in the countries implementing and not implementing a universal BCG vaccination was 1.147 and 1.221, respectively. The mean growth factor for the deaths (*f_d_*) was 1.120 and 1.211.

The results of the PCA analysis showed that the countries implementing a BCG universal vaccination policy were again associated with smaller growth factors for the cases (*f_c_*), but this time the effect was not significant (*p* = .2665, *N*= 110). It is worth noting that this outcome possibly stemmed from the fact that the BCG vaccination policy was significantly confounded with PC1 (*p* = 3.62×10^−11^, Kruskal-Wallis test).

Regarding *f_d_*, the model included 57 countries that met the inclusion criteria. The country’s BCG policy was the only significant predictor (*p* = .0114) and was associated with smaller growth factors. The results are summarized in Table 7.

**Table 7.** Models of growth factor of cumulative cases and deaths with the PCA components.

| Variable | β-Coefficient | $p$-value | Unique Variance |
| --- | --- | --- | --- |
| *Cases<sub>pm</sub>* |  |  |  |
| *BCG Policy* | -0.0870 | .2665 | 0.24% |
| *TB Incidence* | -0.0199 | .7952 | 0% |
| *Total Population* | 0.2723 | .0490 | 2.77% |
| *PC1* | 0.5353 | $4.60 \times 10^{-9}$ | 27.78% |
| *PC2* | 0.2885 | .0299 | 3.57% |
| *PC3* | -0.1273 | .0452 | 2.90% |
| *Deaths<sub>pm</sub>* |  |  |  |
| *BCG Policy* | -0.3979 | .0114 | 10.38% |
| *TB Incidence* | -0.1697 | .2436 | 0.76% |
| *Total Population* | 0.1701 | .5214 | 0% |
| *PC1* | 0.0957 | .5585 | 0% |
| *PC2* | 0.3847 | .1206 | 2.85% |
| *PC3* | 0.1595 | .1587 | 2.01% |

Incidentally, the differences between the two sets of models (e.g., *β* = 0.5353 and *β* = 0.0957 for PC1) might be due to the fact the model of deaths_pm_ include many fewer countries than the model of cases_pm_ (*n* = 57 and *n* = 110, respectively).

## Discussion

This study examined the potential effects of countries’ BCG vaccination policy and TB incidence on the occurrence of COVID-19 recorded cases and deaths (as of April the 26^th^ 2020). The results highlight that both variables exhibit an effect on the number of the recorded cases and deaths per one million population related to the COVID-19. Notably, the effects of the two variables appear to be additive (≈ 5% to 16% of total unique variance explained; Tables 1, 2, and 3). These effects remained largely significant even after controlling for several potential confounders.

A potential concern of the above analyses was that we used data that were not corrected by the onset of the spread of the virus. Thus, we have also investigated the putative protective effects of the BCG vaccine and TB incidence in a fixed time window (15 days) whose beginning is determined by a specific threshold in the number of cases (e.g., one case per one million population or 100 absolute cumulative cases). We have run this analysis to estimate the effects of the dependent variables of interest on the growth factor linked to the cases and deaths at the initial stage of the pandemic in each country. Overall, the results confirm the effect of the BCG vaccine. The only exception is the PCA analysis for the cases in the second growth-factor analysis (*p* = .2665). On the other hand, the country’s TB incidence seems to have little impact on the growth factors, possibly because this variable may show an effect in later stages of the pandemics, or in delaying the initiation of the growth (See Figure S21), or because its impact on growth factors does not add to the one of BCG policy. Concerning the relationship between BCG policy and TB incidence, and potential contribution of other infectious diseases, such as, rubella, measles, and malaria, see the discussions in “Tuberculosis and other infectious diseases” in Supplementary Text for the details.

Finally, the analysis of the strain results may suggest that the G3− strains of the BCG vaccine were more effective than the G3+ strains. Currently, the role of the type of BCG strain is, however, still uncertain. Our analysis is limited to a simple comparison (t-test) between those countries that have been using G3− strains and those ones that have been using G3− strains. Since the type of strained used is severely confounded with the BCG policy adopted by the countries (i.e. most countries that discontinued mandatory BCG policy adapted G3+ strains), it is technically hard to dissociate the effect of BCG strains from BCG policy. For the details, see discussions on “BCG strains” in Supplementary Text.

BCG, originally developed against tuberculosis, is hypothesized to develop ‘frontline’ immunity, training it to respond non-specifically to certain viruses with greater intensity^7,3,4^. This idea is supported by clinical and epidemiological studies, which showed that BCG appeared to lower overall mortality in children^5,2,6^. BCG, which can remain alive in the human skin for several months, triggers not only specific memory B and T cells, but also stimulates the innate blood cells for a prolonged period^7^. In a randomized placebo-controlled study, it was shown that BCG vaccination protects against experimental infection with a weakened form of the yellow fever virus^7,4^. By showing a strong association of BCG vaccination program with COVID-19-related risks, our results are in line with the idea that BCG vaccination provides non-specific protection against COVID-19.

Should the exposition to tuberculosis (either via the vaccination or disease) exert a protective effect against the novel COVID-19, the advantages could be substantial. For example, the results of our growth factor analysis show a moderate mean difference in recorded cases between the countries that implement a universal BCG vaccination policy and the ones that do not (see Tables 4 and 5). Nevertheless, a small difference in an exponential function can lead to large effects. Consider the following counterfactual comparison based on our estimated impacts from the growth factor analysis: Assuming *N_0_* = 100 in two groups at day 0, our results suggest that after 15 days countries currently administering the vaccine would see about 780 cases (mean growth factor = 1.147). All else equal, countries with no universal vaccination policy would see about 2,000 cases (mean growth factor = 1.221). Furthermore, in one month, the cases would be about 6,000 and 40,000, respectively. Because of the underlying exponential trend of the disease spread, a seemingly small mean difference in the growth factor (0.074) can translate to a massive impact on the number of total cases/deaths in a relatively short amount of time.

We must discuss the limitation to interpret the present findings. A serious limitation is that we only conducted an ecological study, which does not exclude the effect of unknown confounding factors. We cannot determine the definitive causality link. Future clinical trials that manipulate the potential confounding variables can test causality. Individual-level epidemiological studies that examine individual history of BCG vaccination and TB infection would also contribute to testing this hypothesis.

## Conclusions

Overall, these findings corroborate the hypothesis that BCG vaccination and exposure to tuberculosis may induce a non-specific protection against the novel SARS-CoV-2 infection, while limitations of ecological study need to be recognized. Due to the potential public-health benefits, our results indicate that this hypothesis deserves further attention and should not be hastily dismissed.

## Data Availability

The data are available in the supplemental materials. The sources of the data are retrievable from the reference list.

## Acknowledgement

We thank Yoko Kagami, and Harumi Mitsuya for their assistance in inputting the data and making Figures, Jun Sato, Mamoru Tejima for letting us know the existence of potential correlation and for valuable discussions, Colleen Bailey for providing us data of potentially confounding variables, Japan BCG for providing us information on BCG Tokyo, and Hideo Hagihara, Tomoyuki Murano, Hironori Funabiki, Shinichi Nakagawa, Takashi Inoue, Hiroyuki Morita and many other twitter users for useful discussions.

## Supplementary Discussions

### Tuberculosis and other infectious diseases

Because BCG is an attenuated version of the bacterium causing TB, it is biologically plausible to assume that experience of TB infection itself may have a protective effect against COVID-19. About 80% of people in many Asian and African countries test positive, while 5–10% of people in the United States population test positive in the tuberculin test^44^. These numbers are consistent with our findings: high burden countries, many of which are in Africa and Asia, show low COVID-19 indices, even at this stage (April 26^th^). There are 53 high TB burden countries and regions, and, surprisingly, in the top 50 countries/regions among 142 in terms of the number of deaths per 1M population, we found only three high TB burden countries (Table S1). High TB burden countries are mostly countries with low income, which is, in fact, counter-intuitive, considering that low-income countries generally suffer from low hygiene and low medical care.

Obviously, the decision of a country to discontinue BCG vaccination depends on TB prevalence, and so it is difficult to dissociate the effects of the two variables. Nonetheless, we have found additive effects of BCG group and TB prevalence. Also, there is the effect of the BCG vaccination occurs in low-TB-prevalence countries (Figure 2), which is consistent with the idea that TB infection and BCG vaccination may have similar protective effects against COVID-19.

It is worth noting that a high TB burden is likely to be associated with a lower hygiene and a high prevalence of other infectious diseases, such as malaria, dengue fever, yellow fever, rubella, and measles. In fact, the prevalence of rubella, measles, and malaria are all weakly and negatively correlated with low COVID-19 indices (Table S6; Figures S15, S16, and S17, respectively), as is the prevalence of TB (Figure 2). It is possible that these diseases, which are common in the regions where the TB burden is high, and/or some of the vaccines and drugs administered to fight these diseases may induce improvement in non-specific immunity or inhibit replication or function of SARS-CoV-2.

Regarding ethnic genetic background, which is associated with a high TB burden in African and Asian countries, see the discussion in the section, “Ethnic genetic background.”

### BCG Strains

BCG was derived from Mycobacterium bovis, a bacterium causing tuberculosis in cattle, and was attenuated after 230 passages over a period from 1908 to 1921^25^. Throughout these passages, many variations of strains were generated. Currently, several major strains are usually employed, including BCG Tokyo, Russia, Pasteur, and Denmark. While these strains are derived from the same bacterium, they have substantial differences in their characteristics, including genomic sequences^25^, the influence on immunological responses^45^, and their effectiveness in protection against TB^46^. Regarding the differences in genomic sequences, Zhang et al. conducted a comparative genomic analysis among several BCG strains and strains of M. tuberculosis. They found that BCG strains have progressively been suffered from deletions in the genomic regions containing T-cell antigen epitopes, and that BCG strains have strain-specific regions of deletions. The strains most commonly in use, such as BCG Denmark and Pasteur, have the largest number of regions of deletions, while BCG Russia and Tokyo have the least. Some evidence suggests that BCG Tokyo induces a stronger immune response than other strains^27,47^. It has been proposed that BCG Tokyo strain, which contains the largest number of T-cell epitopes and is the only strain with the same number of epitopes as the first BCG vaccine strain in 1921, may be the best candidate strain for the development of a better vaccine^25^. In the present study, based on the work of Zhang et al.^25^, we have classified the BCG strains into two categories – *G3+* and *G3−* strains – which either still have or have lost 28 “Group 3” epitopes, all of which are located in RD2 (Region of Deletion 2). Our results suggest that G3− group (Tokyo, Russia and Moreau), has a higher protective effect on COVID-19 than G3+ group (Denmark, Pasteur) (Figure 1-4). This is consistent with the idea that the strains that have fewer genomic deletions and retain more original epitopes may be more effective against COVID-19. As can be seen in the Table S1, there are a number of countries that lack the information about the BCG strain used. This information is thus needed for more detailed studies evaluating the potential functional differences of BCG strains in their efficacy against COVID-19.

### Life expectancy, population of ages 65 and above, number of beds per population and median income

The highly significant positive correlations between life expectancy / median income and COVID-19 indices (Table S4; Figure S1: Life expectancy; Figure S2: Median household income) are among the unique and counter-intuitive characteristics of this disease. In fact, one might actually expect a negative correlation since wealthy long-life-expectancy countries usually benefit from a better hygiene and medical care compared to poor countries. In the same vein, there are positive correlations also between the number of hospital beds and COVID-19 indices (Figure S7). In addition, negative correlations between absolute income and mortality in general have been reported^48,49^. In subpopulation analyses, by adjusting each of these variables among BCG groups, the effects of BCG group on COVID-19 indices remained significant (Figure S1: Life expectancy; Figure S2: Median household income; Figure S7: Hospital beds per 1000 people). Therefore, it is obvious that long life expectancy, high income, and high-quality medical care themselves cannot solely account for the between-country differences in COVID-19 cases/deaths.

There are at least three factors that may account for this positive correlation, other than TB incidence. First, older adults seem to be at higher risk of developing severe complications from COVID-19. In the USA, 31% of cases, 45% of hospitalizations, 53% of ICU admissions, and 80% of deaths associated with COVID-19 have occurred in older adults (aged ≥65 years)^50^. Countries with longer mean life expectancy host a higher number of older adults, who tend to be more vulnerable to COVID-19. Second, wealthy countries usually report a higher proportion of people who suffer from diseases associated with obesity and metabolic syndrome (e.g., cardiovascular diseases and Type 2 diabetes). These conditions appear to be major risk factors that increase the mortality from COVID-19. Third, social factors, such as population density and travels, may be more common in rich countries than less wealthy ones. Indeed, these factors are highly correlated with each other and with COVID-19 indices (Table S4) and may, to some extent, contribute to increasing the spread of the virus. (We will take up this point below in more detail.)

We conducted sub-population analyses with each one of these factors and, after adjusting each variable among BCG groups, the effects of BCG group on the COVID-19 indices remain significant (Figure 4: Urban Population as a fraction of total population; Figure S5: International arrivals; Figure S6: Prevalence of overweight adults; Figure S9: Total urban population; Figure S10: Population density). These results suggest that each one of these factors cannot solely account for the effect of BCG group.

### Frequency of overweight individuals

The frequency of overweight individuals is moderately correlated with COVID-19 indices in the worldwide analyses (Table S4; Figure S6).This outcome is consistent with the idea that obesity and its associated diseases, such as diabetes and cardiovascular conditions, increase the risk of contracting and dying of COVID-19^35–37^. However, this relationship is not observed in the European countries (Table S1). Therefore, this variable alone does not seem to be able to explain the pronounced effects of BCG policy / strain groups on the COVID-19 indices in Europe. The observed correlation between the rate of overweight individuals and the COVID-19 indices may be at least partly due to the differences in the COVID-19 indices among world regions. In Europe and the US, COVID-19 is spread more widely and obesity is more common than most other regions, such as Asia and Africa. In our multiple linear regression model, obesity does not exert a statistically significant effect. Also, after adjusting this variable among BCG groups, the effect of BCG group on COVID-19-related cases and deaths remains significant (Figure S6). However, the impact of obesity on the COVID-19 cases and deaths may have been clouded by its collinearity with other variables (Urbanization and Age_65_). In fact, it is likely that obesity does contribute to reducing immune response to COVID-19 infection to some extent. Nonetheless, it may be hard to detect small effects with a between-country design (as in this study).

### Population and traveling activity

Social distancing is believed to affect the spread of COVID-19. We evaluated population-related variables, namely population, land area, population density, and fraction of urban population (for the details, see Table 1). The fraction of urban population, but not total urban population, population density, and land area, is strongly correlated with COVID-19 indices (Table S4; Figure 4: Urban population as a fraction of total population; Figure S9: Total urban population; Figure S10: Population density; Figure S11: Land area). Since the percentage of urban population of most countries that do not currently adopt a universal BCG vaccination policy is 75% or higher, we have evaluated the effect of BCG groups in those countries above the 75% threshold. The effect of the BCG policy remains highly significant even in this subgroup of countries (Figure 4). This result suggests that this factor cannot solely account for the effect of BCG groups in the COVID-19 indices.

How frequently people travel may be another confounding factor. The number of international arrivals is significantly correlated with the country’s COVID-19 cases and deaths (Table S4; Figure S5). However, when we restrict the samples to those countries with a high number of international arrivals to reduce the correlation, the effect of the BCG vaccination policy remains highly significant (Figure S5). Also, presumably, countries that have higher number of travelers from China may be associated with higher COVID-19 indices. However, according to a survey^4^, the top 10 outbound destinations of Chinese travelers are Japan, Thailand, South Korea, Indonesia, Singapore, Malaysia, Australia, UK, New Zealand, and the Maldives. Except for the UK, these countries have recorded extremely low rates of COVID-19 cases and deaths (four or lower deaths per one million population, as of April 26^th^). Thus, travelers from China are unlikely to be a serious confounding variable in our investigation.

### The number of tests for SARS-CoV-2 infection

Political and economic variations between countries may significantly influence COVID-19 testing rates^22^. Hensel et al^18^ reported a highly significant correlation between the number of cases per million and the number of tests for SARS-CoV-2 infection per million. They further report that classifying countries into “High Testing” and “Low Testing” groups makes the significant differences of COVID-19 indices between BCG policy groups disappear. In our study, we also find significant correlations between the number of tests and COVID-19 indices, but the differences between BCG groups remain highly significant, even after controlling for the number of tests per million population (Figure S3). The failure of Hensel et al in detecting the effect of BCG group could be simply explained by the fact that they did not compare incidences between the “Current universal BCG policy” group and the “Universal BCG policy in the past” group. That is, if those groups had been compared, a significant effect would have been detected. Further, they did not find differences in percent mortality per total COVID-19 cases among BCG policy groups, except for a significant difference between “Current universal BCG policy” group and “Never BCG policy” group in “High Testing”. This is consistent with a highly significant correlation between cases and deaths per 1 million population (r = 0.89).

Another possible explanation stems from the prevalence of a large number of untested cases, especially in low income and high TB burden countries. If so, the actual number of infected people may be larger in these countries. Such underestimation, if any, would be reflected in high percentage positive cases per test, since highly symptomatic people are likely to be tested in most of the countries, regardless of their economic wealth, and this tendency could be more pronounced in the countries with low-income/low-quality health care systems due to limited testing resources. However, we did not find a significant negative correlation between the percentage of positive cases per test and median household income (Table S4; Figure S4). Rather, the opposite tendency occurs: high positive correlations were found. Relatively low percentage positive cases per test in low-income countries, especially those in Africa, suggest that the number of total cases in those countries may not be necessarily underestimated. Moreover, regardless of the countries within high or low range of percentage positive cases per test, the effect of BCG groups remained highly significant (Figure S4). These results demonstrate that the variability of the number of tests conducted cannot solely account for the effect of BCG group on COVID-19 indices.

### Prevalence of smoking tobacco

Smoking, by its association with respiratory diseases, may also have negative impacts on the immune system. This, in turn, would make smokers more vulnerable to infectious diseases^52^, including COVID-19^53^. Studies suggest that smokers were more likely to (i) have severe symptoms of COVID-19, (ii) be admitted to an ICU, (iii) need mechanical ventilation, and (iv) die compared to non-smokers^37,54^. In our study, as expected, we find moderate positive correlations between the prevalence of smoking any tobacco product among persons aged >= 15 years and COVID-19 indices (Table S4; Figure S8). BCG group N shows a tendency of middle to high prevalence of smokers (Figure S8). However, significant effects of BCG group remain even in a subgroup of the countries of medium level of smoking prevalence (15-30 % of total population; Figure S8), suggesting that it may not be a serious confounding factor in the effect of BCG group on COVID-19 indices.

### Temperature and absolute humidity

Studies have revealed the effect of temperature and humidity on respiratory virus stability and transmission rates and on host intrinsic, innate, and adaptive immune responses to viral infections in the respiratory tract in general^55^. COVID-19 risk has also been associated with temperature and humidity^33,34,56,57^. Absolute humidity was associated with the local exponential growth of COVID-19 across provinces in China and other affected countries^57^, while, in another study, no significant association between COVID-19 incidence and absolute humidity was observed^56^. We find a moderate and weak negative correlation between temperature / absolute humidity and COVID-19 indices, respectively (Table S4; Figure S12: Absolute humidity; Figure S13: Temperature). BCG group Y tends to have a higher temperature, and to circumvent the confounding effect, we compared BCG groups in the countries with intermediate temperature rage (0-15°C), but the effects of BCG remained significant (Figure S13). Likewise, BCG Group N tends to have relatively low absolute humidity and we analogously evaluated the effect of BCG groups on COVID-19 indices among the countries that have 10 or less absolute humidity (g/m^3^). The effect of BCG groups remained significant (Figure S12) in this subsample as well. These results suggest that neither temperature nor absolute humidity can explain the effects of BCG groups we observe.

### Community mobility change / social distancing

Studies have suggested the potential of a lockdown or social distancing to slow down the spread of COVID-19^58–62^ and the strength of social distancing could be a confounding factor in the apparent effect of BCG groups. To evaluate this possibility, we obtained COVID-19 community mobility reports by Google^63^. We used the averages of mobility change (%) from baseline mobility in three indices (“retail & recreation”, “transit stations”, and “workplaces”) in each country. We calculated (1) averages of the data during the period between March 1^st^ to April 15^th^ and (2) the ones during 15 days from 14 days before the 100^th^ case was detected until the 100^th^ case day.

Unexpectedly, we found negative correlations between mobility change (average of March and April) and COVID-19 indices (Table S4; Figure S14). The countries that are experiencing high COVID-19 burden may tend to employ strong lockdown or social distancing measures and the negative correlations could be due to this tendency. We compared BCG groups in the countries with intermediate mobility change (from −30% to −50%), and found that the effects of BCG remain significant (Figure S14). We also included the mobility change (the one before 100^th^ case) in our growth analyses, which did not make any significant changes to our results. These results suggest that the differences in community mobility change or social distancing among countries does not explain in our observed effects of BCG groups.

### Ethnic genetic background

African and Asian countries tend to have lower COVID-19 indices compared to the countries in Europe and North America (Table S1). This could be potentially due to some biological differences of ethnic genetic background. Genetic variations of the genes, such as the ACE2, TMPRSS2, and HLA, are among the candidates that affect susceptibility to COVID-19^64,65^. In the case of the USA, Asian, Latino and Whites are similar in terms of mortality rate per 100,000 residents of each group (16, 15.9, and 14.3 per 100K residents, respectively), while that of African Americans is much higher (37.2 per 100K residents) ^66^. Indirect standardization of mortality data shows that Black, Asian and minority ethnic groups in the United Kingdom are at an increased risk of death from COVID-19 compared to Whites^67^. The higher mortality from COVID-19 in these races are considered to be partly due to low income^67^. These data fail to support the hypothesis of a protective effect arising from the unique genetic and ethnic background of people in African and Asian countries. This seemingly contradictory relationship between low life expectancy/low income and low COVID-19 indices in many African and some of the Asian countries might be better explained by innate immunity improved by high prevalence of TB in these countries, and BCG vaccination.

### Duration of BCG vaccination

The duration (years) over which a country had mandatory BCG vaccination policy may matter^11^. Consistent with the results with the previous study, we can see a weak negative correlation between the BCG duration and COVID-19 indices in BCG N countries, but we did not find clear correlation in BCG Y countries (Figure S18). This difference between the previous study^11^ and ours might be due to the fact that we included the data from “low-income countries”, which were excluded in the previous study. It is noteworthy that there are virtually no high COVID-19 burden countries in high TB burden countries, defined by 100 incidents per 100K populations, even with short BCG duration (Figure S18). Miller et al ^11^ discussed that this might be due to “under-reporting” of the cases/deaths, which is unlikely to be the case (see the section, “The number of tests for SARS-CoV-2 infection”). Moreover, our multiple linear regression analyses suggest that TB incidence has an effect on reducing COVID-19 indices. Therefore, the interpretation would be more plausible that low COVID-19 burden in the countries with short BCG length would be mostly due to their high burden of TB and/or other infectious diseases, that may have a protective effect against COVID-19.

### Other points

In this study, while we have assessed 19 potentially confounding factors in total, we recognize that other factor(s) may underlie this apparent BCG/TB effect. Consider, for instance, the differences between the strains of SARS-CoV-2 dominant in each country/region^68^. As far as we know, however, no studies have found clear evidence for functional differences among different strains of this virus, and this possibility needs to be evaluated in the future.

Another possible factor could be the languages used in each country. At the time of the outbreak of Severe Acute Respiratory Syndrome (SARS) in 2003, the difference in droplet production among languages was suggested to affect the efficiency of transmission of SARS^69^. The language in China, where the outbreak occurred, has an aspiration/non-aspiration pronunciation system, which could produce more droplets^69^. English, but not Japanese, uses a similar pronunciation system and this kind of difference could underlie the difference in the transmission of SARS-CoV-2. However, there have been few high COVID-19 burden regions that use the Chinese language, except for Wuhan, and it is unlikely that this factor can solely explain the apparent effect of BCG/TB.

Also, custom or policy for wearing masks may matter, but the differences between western and eastern Europe and the low COVID-19 prevalence in African countries may argue against the idea that this is a major confounding factor underlying the apparent effect of BCG/TB.

In each sub-population analysis that evaluated the effect of confounding variable, we used the countries that fell within a window, in which average values of the variable of each BCG group become approximately same. In this series of the analyses, many African and Asian countries tended to get dropped, mainly because they are outliers regarding life expectancy, median income, international transits, and number of tests. Many of these African and Asian belong to currently BCG Y and high TB burden group, which we hypothesize to be most protective. The observed impacts, after dropping these countries that have low COVID-19 cases/deaths, therefore, is a conservative estimate of our hypothesized impact. The fact that the effect of BCG policy remains significant even after this exclusion suggests that the effects are robust.

Various other potential confounders are not assessed in our study. Examples include the consumption of nutrition, such as vitamin D and antioxidants, and the use of drugs, such as ivermectin. Also, not a single factor but combinations of multiple factors could be causing such apparent BCG/TB effect. This is a limitation inherent in the research design based on cross-country comparisons, and future studies are needed to test this hypothesis.

Supplementary Figures

**Figure S1.**
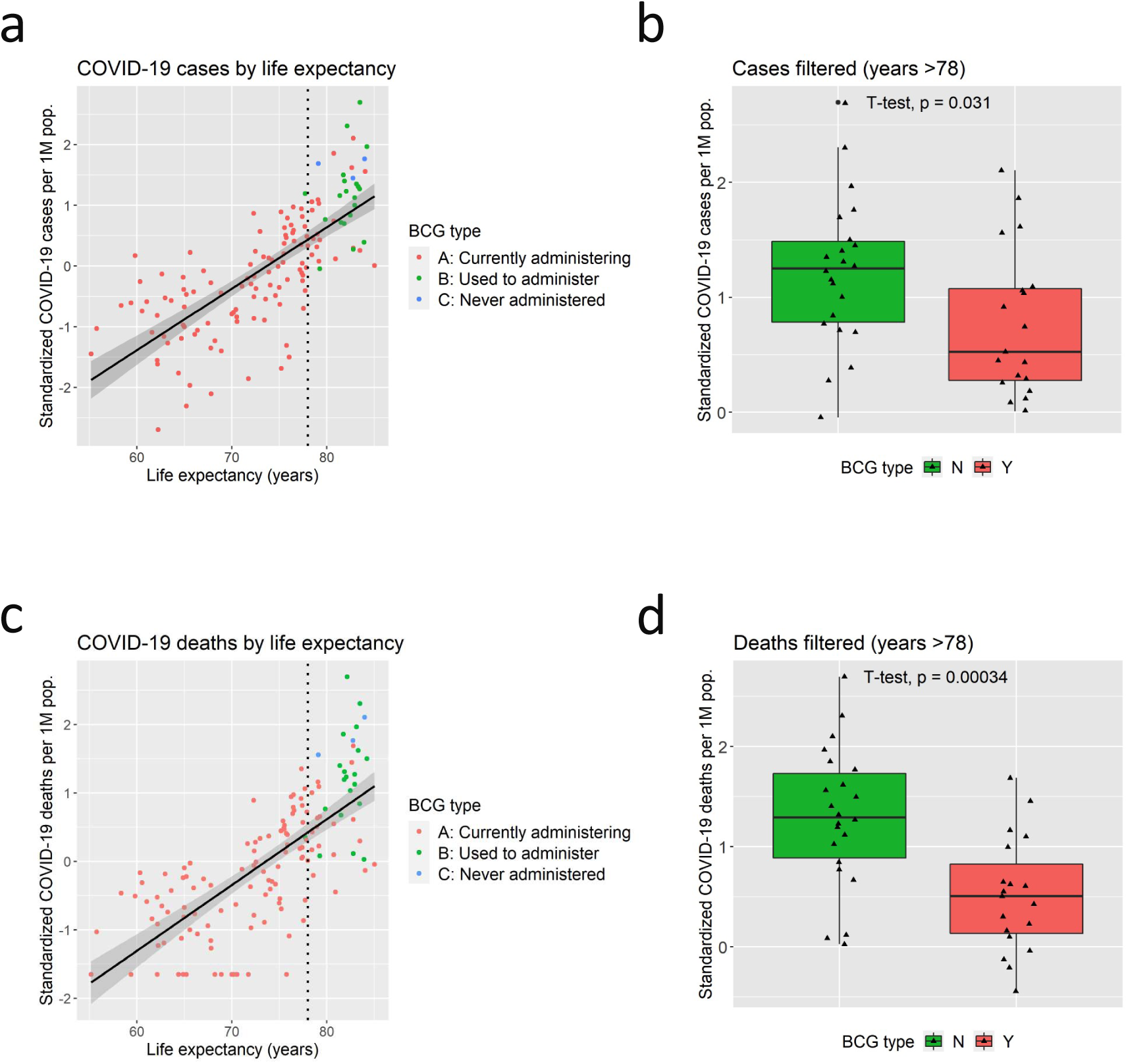
Correlations between life expectancy and COVID-19: a. Scatter plot of total cases per one million population by life expectancy. The vertical dotted line indicates the threshold of 78 years, that is subsequently used for the analyses shown in the panel b. b. The boxplot of total cases per one million population sorted by BCG Group in countries with life expectancy higher than 78 years. Group N show a significantly higher rate of cases of COVID-19 compared to Group Y. c. Scatter plot of total deaths per one million population by life expectancy. d. The boxplot of total deaths per one million population sorted by BCG Group in countries with life expectancy higher than 78 years. Group N show a significantly higher rate of deaths of COVID-19 compared to Group A.

**Figure S2.**
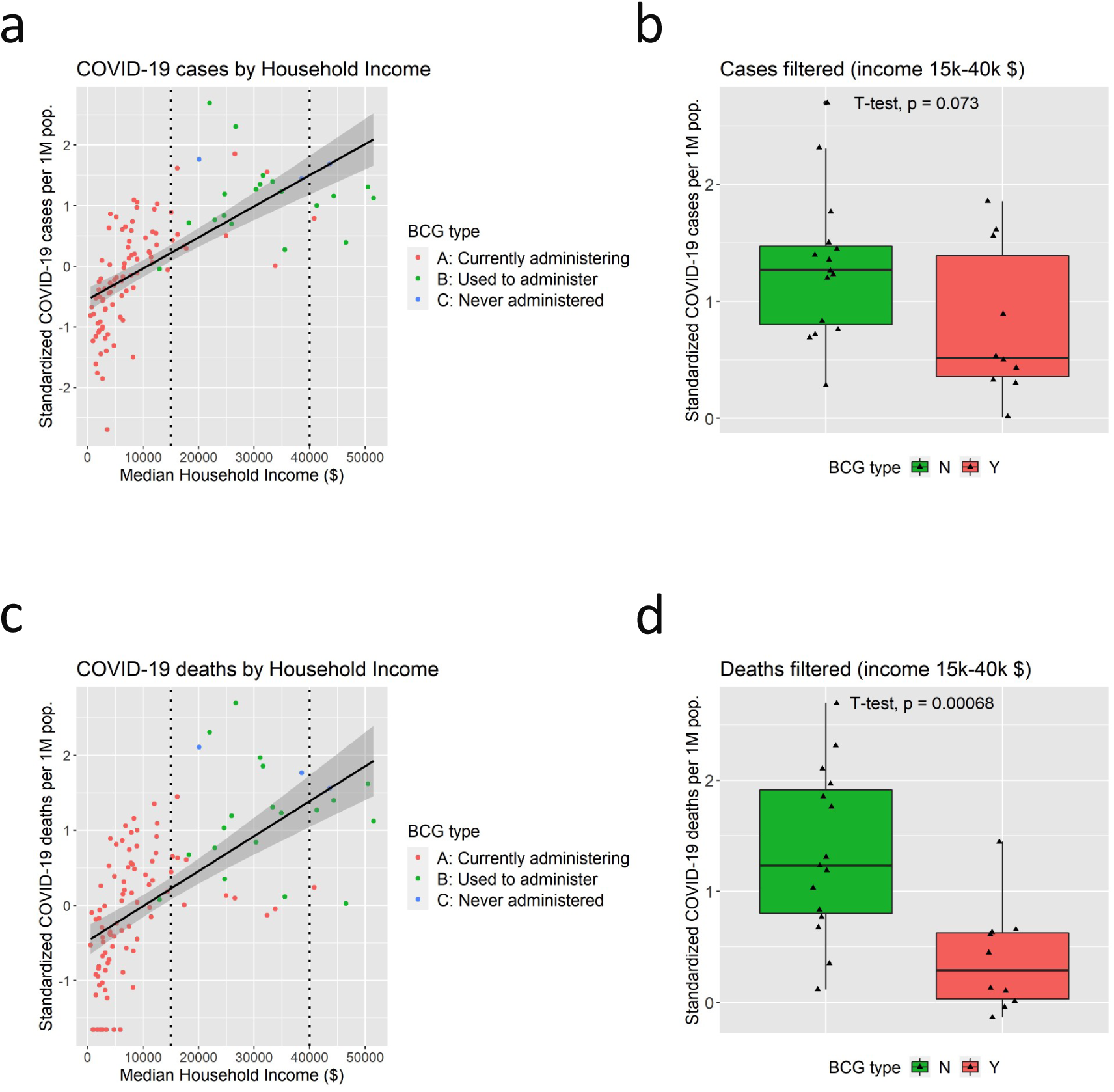
Correlations between Median Household Income and COVID-19: a. Scatter plot of total cases per one million population by median household income ($). The vertical dotted line indicates the threshold of $15,000 and $40,000, that is subsequently used for the analyses shown in the panel b. b. The boxplot of total cases per one million population sorted by BCG Group in countries with median household income between $15,000 and $40,000. Group N show a tendency of higher rate of cases of COVID-19 compared to Group Y. c. Scatter plot of total deaths per one million population by median household income. d. The boxplot of total deaths per one million population sorted by BCG Group in countries with median household income between $15,000 and $40,000. Group N show a significantly higher rate of deaths of COVID-19 compared to Group A.

**Figure S3.**
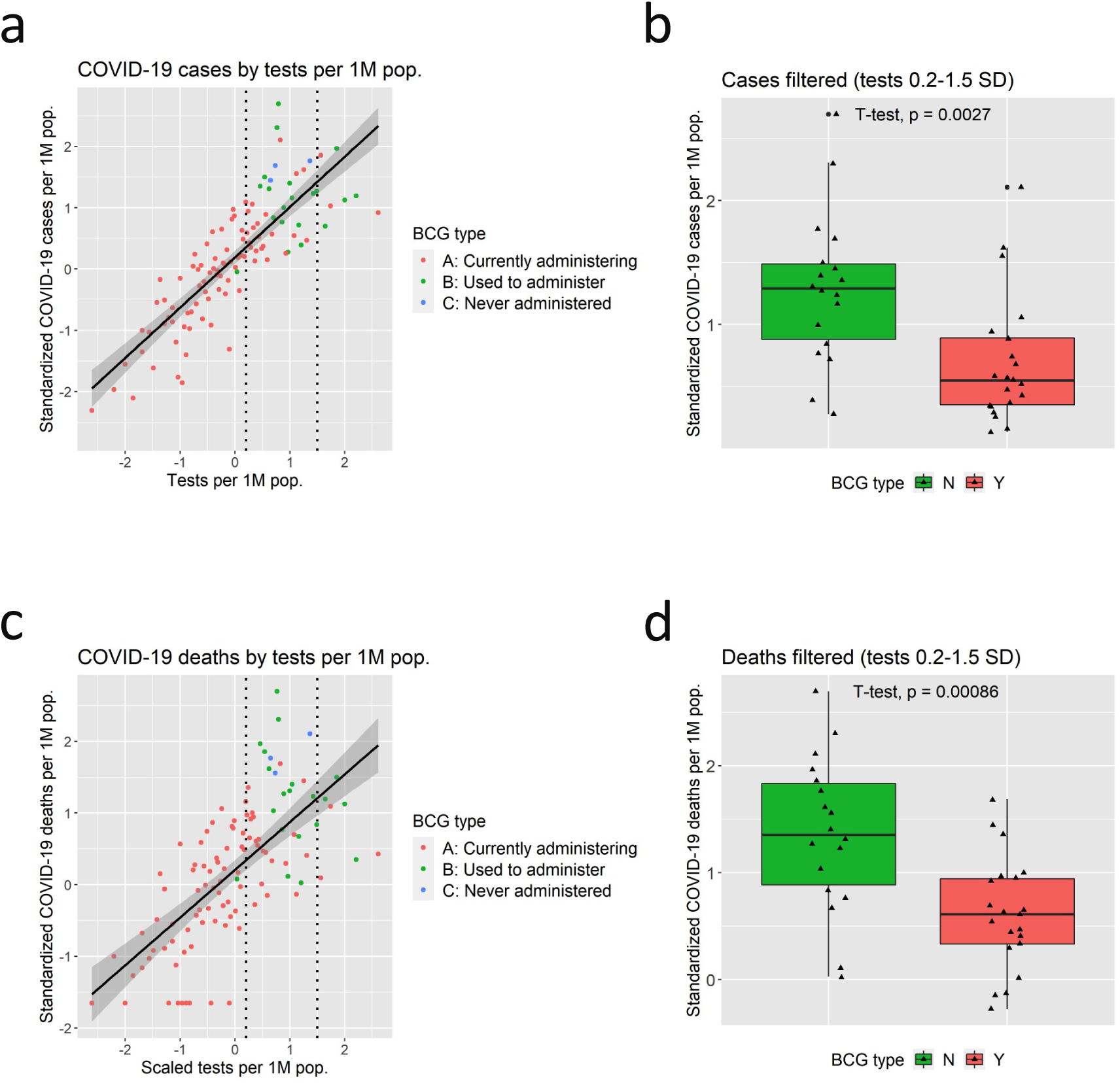
Correlations between Tests per 1 million population and COVID-19: a. Scatter plot of total cases per one million population by the number of tests conducted for SARS-CoV-2 infection per 1 million population. The vertical dotted line indicates the threshold of 0.2 SD and 1.5SD, that is subsequently used for the analyses shown in the panel b. b. The boxplot of total cases per one million population sorted by BCG Group in countries with tests per 1million population between 0.2 SD and 1.5SD. Group N show a significantly higher rate of cases of COVID-19 compared to Group Y. c. Scatter plot of total deaths per one million population by tests per 1million population. d. The boxplot of total deaths per one million population sorted by BCG Group in countries with tests per 1million population between 0.2 SD and 1.5SD. Group N show a significantly higher rate of deaths of COVID-19 compared to Group Y.

**Figure S4.**
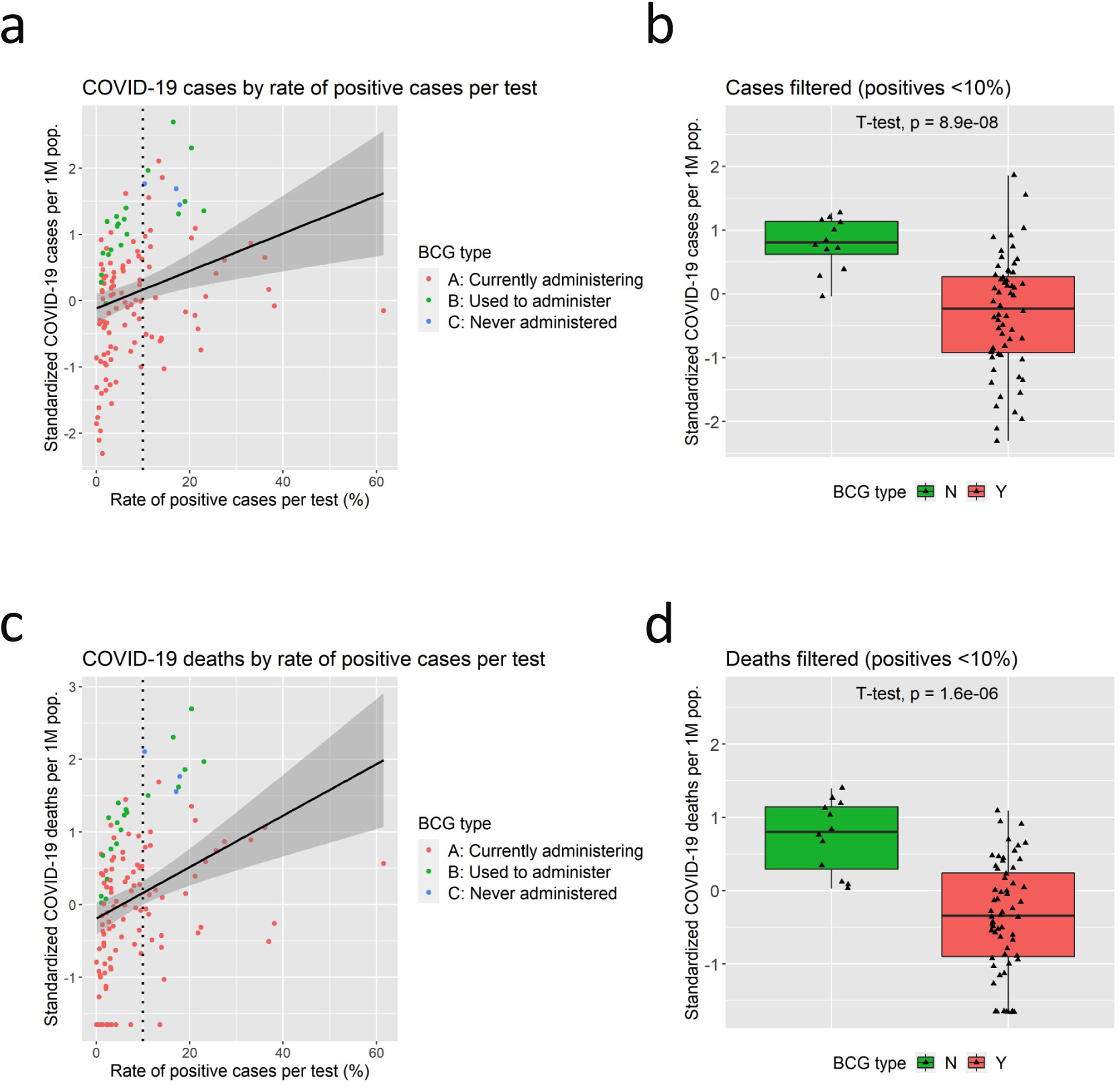
Correlations between rate of positive cases per test and COVID-19: a. Scatter plot of total cases per one million population by rate of positive cases per test (%). The vertical dotted line indicates the threshold of 10%, that is subsequently used for the analyses shown in the panel b. b. The boxplot of total cases per one million population sorted by BCG Group in countries with rate of positive cases per test under 10%. Group N show a significantly higher rate of cases of COVID-19 compared to Group Y. c. Scatter plot of total deaths per one million population by rate of positive cases per test (%). d. The boxplot of total deaths per one million population sorted by BCG Group in countries with rate of positive cases per test under 10%. Group N show a significantly higher rate of deaths of COVID-19 compared to Group Y.

**Figure S5.**
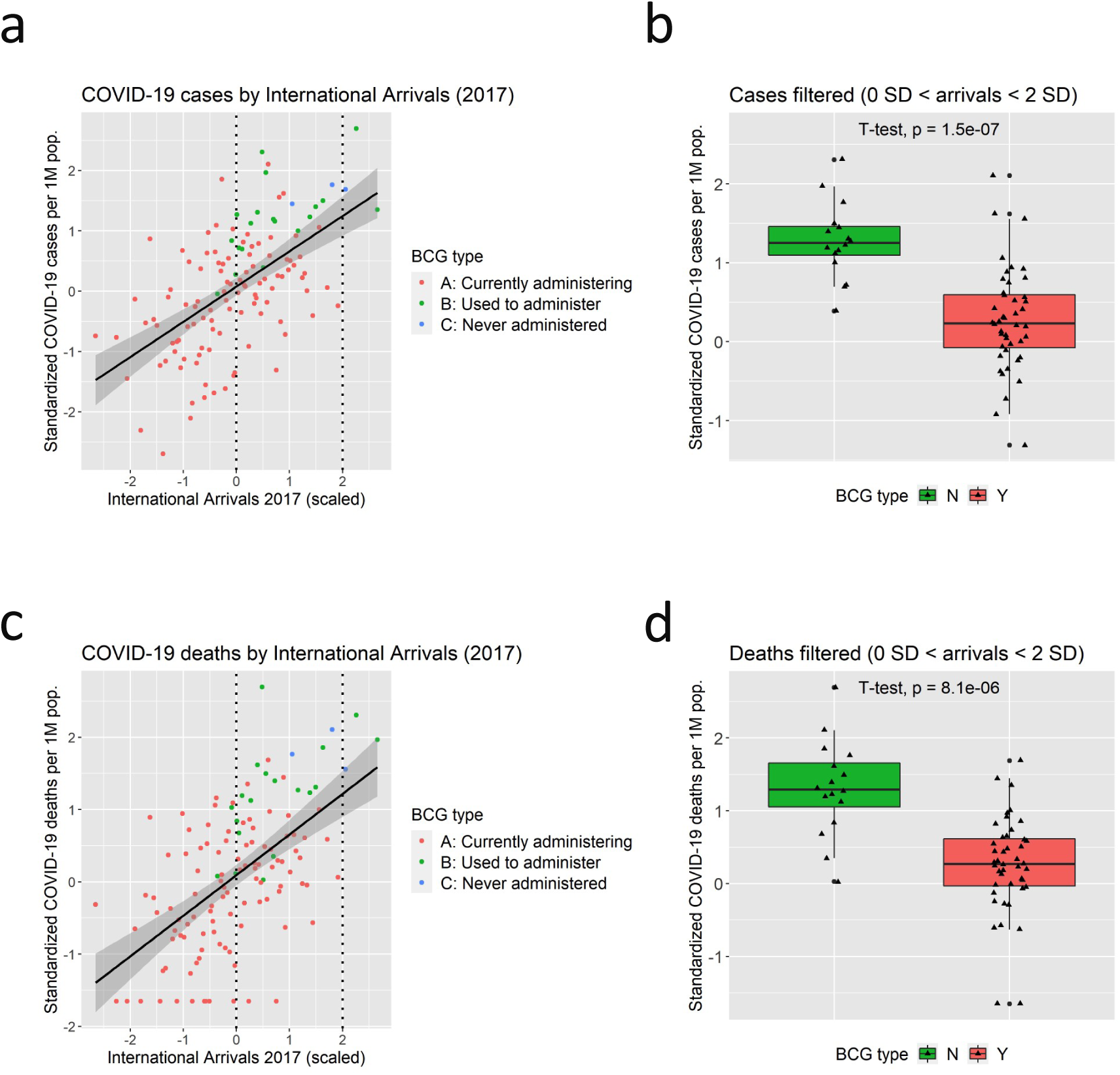
Correlations between International Arrivals and COVID-19: a. Scatter plot of total cases per one million population by the number of international arrivals (scaled). The vertical dotted line indicates the threshold of 0 SD and 2 SD, that is subsequently used for the analyses shown in the panel b. b. The boxplot of total cases per one million population sorted by BCG Group in countries with international arrivals between 0 SD and 2 SD. Group N show a significantly higher rate of cases of COVID-19 compared to Group Y. c. Scatter plot of total deaths per one million population by international arrivals. d. The boxplot of total deaths per one million population sorted by BCG Group in countries with international arrivals between 0 SD and 2 SD. Group N show a significantly higher rate of deaths of COVID-19 compared to Group Y.

**Figure S6.**
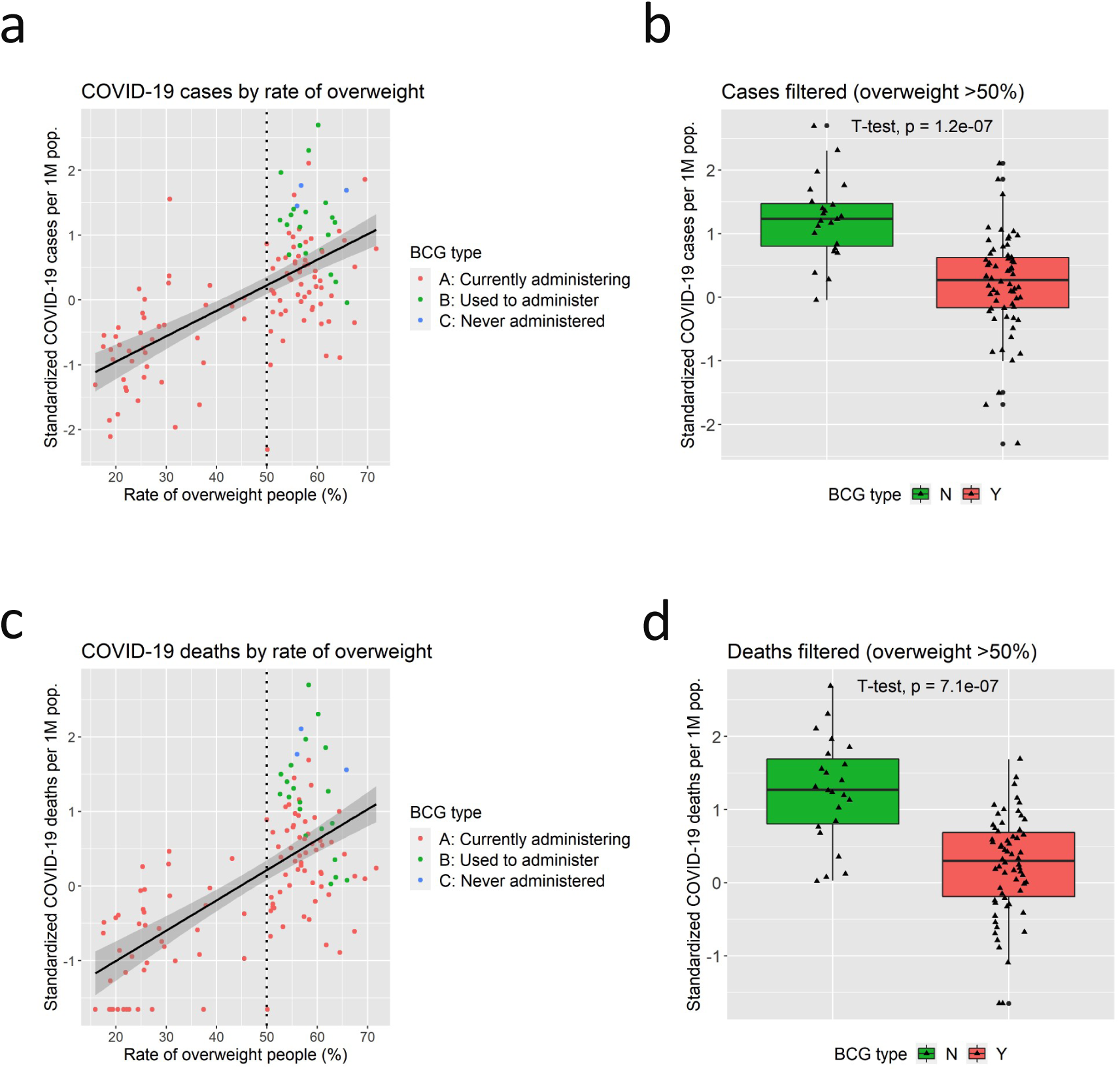
Correlations between Prevalence of Overweight Adults and COVID-19: a. Scatter plot of total cases per one million population by prevalence of overweight adults (%). The vertical dotted line indicates the threshold of 10%, that is subsequently used for the analyses shown in the panel b. b. The boxplot of total cases per one million population sorted by BCG Group in countries with prevalence of overweight adults above 50%. Group N show a significantly higher rate of cases of COVID-19 compared to Group Y. c. Scatter plot of total deaths per one million population by prevalence of overweight adults (%). d. The boxplot of total deaths per one million population sorted by BCG Group in countries with prevalence of overweight adults above 50%. Group N show a significantly higher rate of deaths of COVID-19 compared to Group Y.

**Figure S7.**
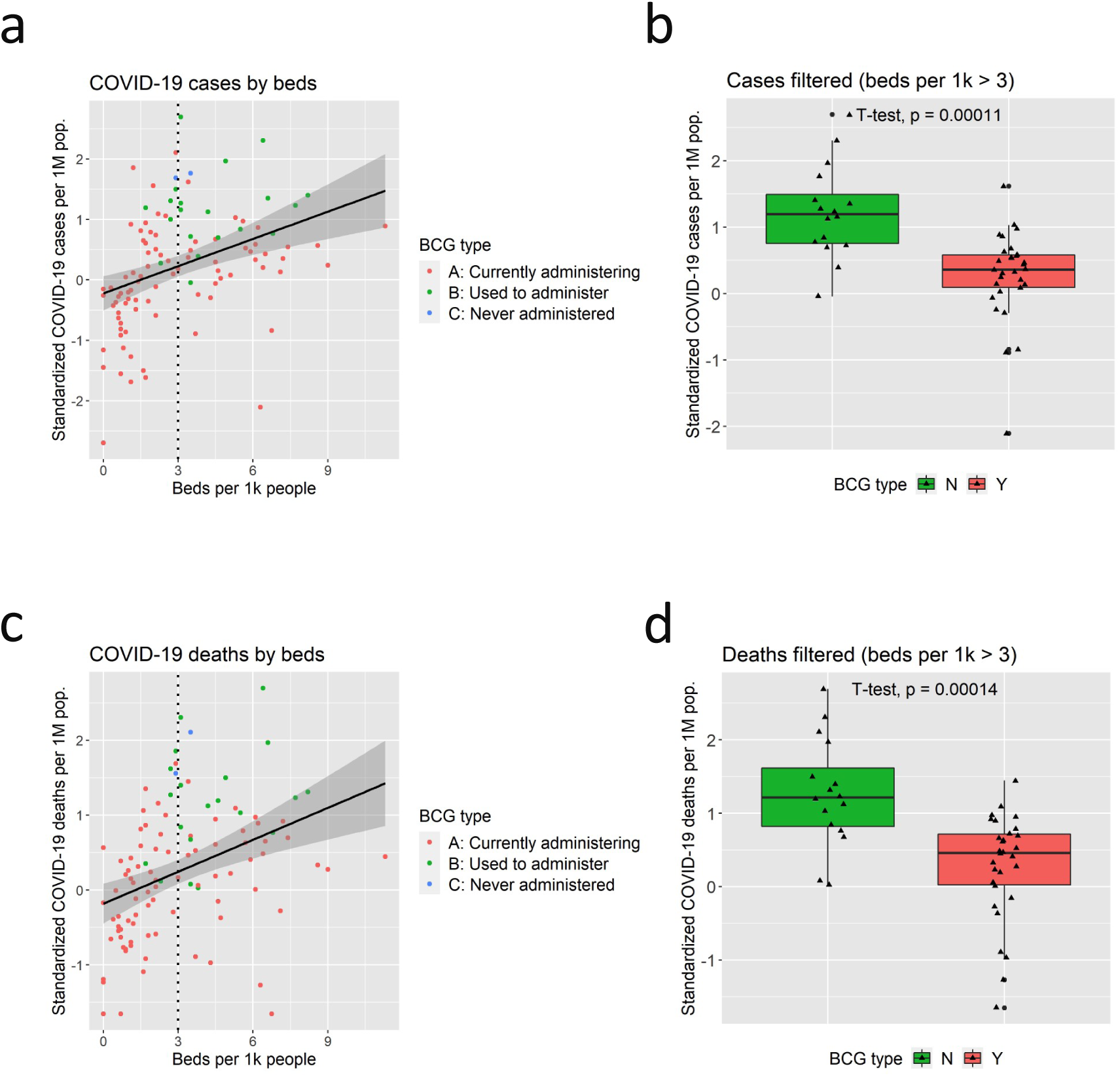
Correlations between hospital beds per 1000 people and COVID-19: a. Scatter plot of total cases per one million population by the number of hospital beds per 1K people. The vertical dotted line indicates the threshold of 10%, that is subsequently used for the analyses shown in the panel b. b. The boxplot of total cases per one million population sorted by BCG Group in countries with hospital beds per 1K people above 3. Group N show a significantly higher rate of cases of COVID-19 compared to Group Y. c. Scatter plot of total deaths per one million population by the number of hospital beds per 1K people. d. The boxplot of total deaths per one million population sorted by BCG Group in countries with hospital beds per 1K people above 3. Group N show a significantly higher rate of deaths of COVID-19 compared to Group Y.

**Figure S8.**
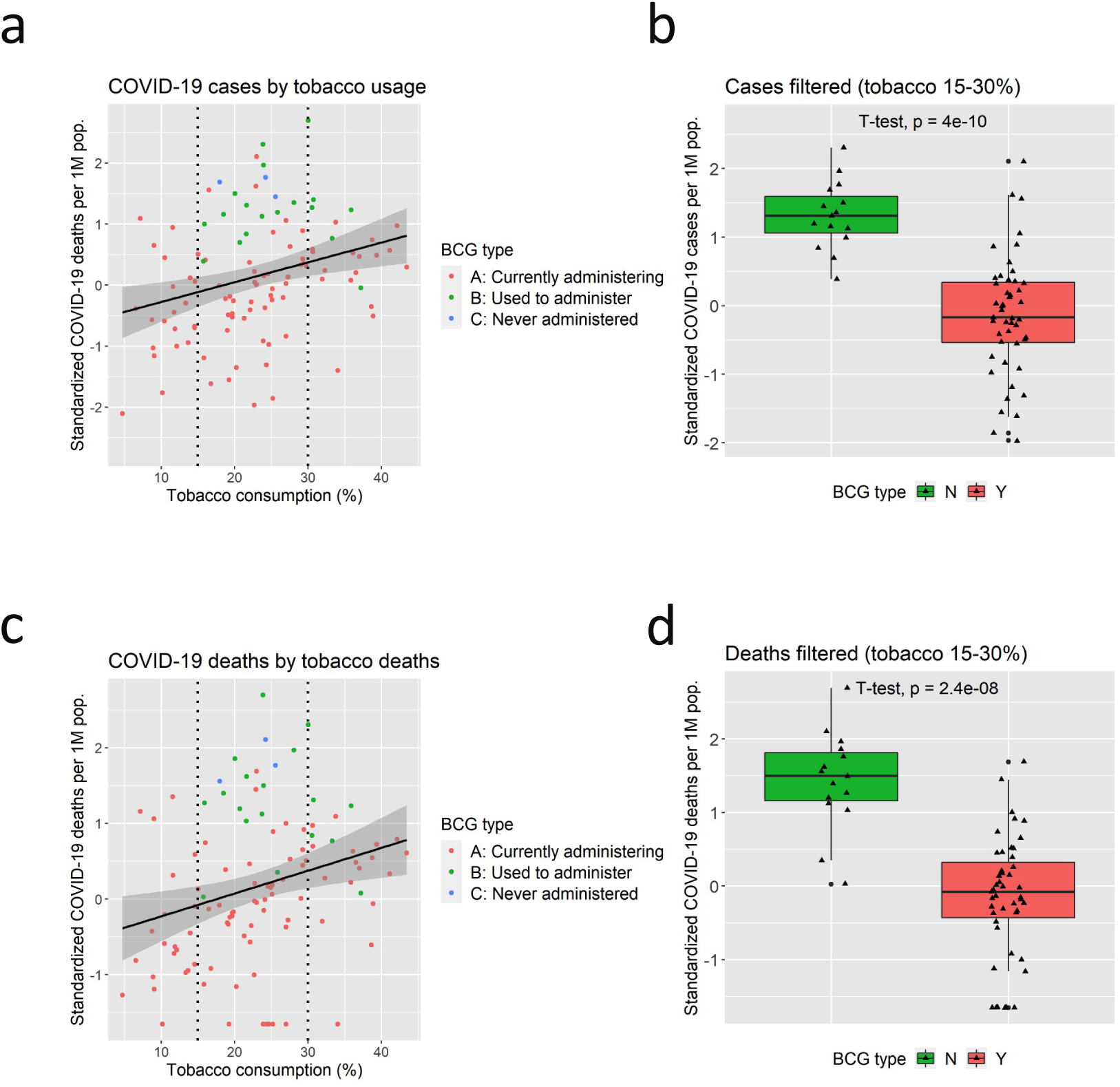
Correlations between prevalence of smoking and COVID-19: a. Scatter plot of total cases per one million population by the percentage of the population aged 15 years and over who currently use any tobacco product (%). The vertical dotted line indicates the threshold of 15 % and 30%, that is subsequently used for the analyses shown in the panel b. b. The boxplot of total cases per one million population sorted by BCG Group in countries with prevalence of smoking between 15 % and 30%. Group N show a significantly higher rate of cases of COVID-19 compared to Group Y. c. Scatter plot of total deaths per one million population by prevalence of smoking. d. The boxplot of total deaths per one million population sorted by BCG Group in countries with prevalence of smoking between 15 % and 30%. Group N show a significantly higher rate of deaths of COVID-19 compared to Group Y.

**Figure S9.**
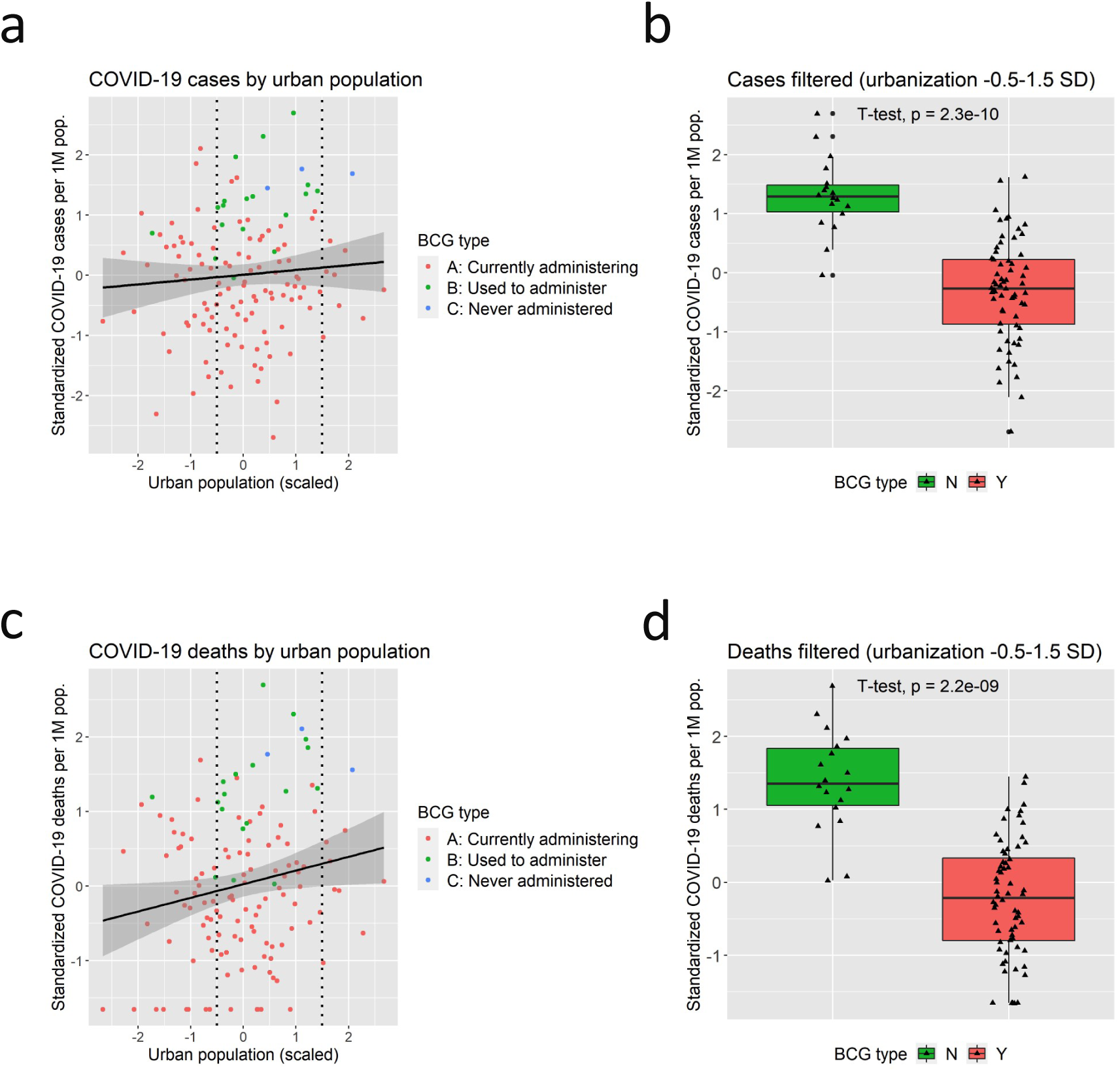
Correlations between urban population and COVID-19: a. Scatter plot of total cases per one million population by total number of people in urban areas (scaled). The vertical dotted line indicates the threshold of 0.5 SD and 1.5 SD, that is subsequently used for the analyses shown in the panel b. b. The boxplot of total cases per one million population sorted by BCG Group in countries with urban population between 0.5 SD and 1.5 SD. Group N show a significantly higher rate of cases of COVID-19 compared to Group Y. c. Scatter plot of total deaths per one million population by urban population. d. The boxplot of total deaths per one million population sorted by BCG Group in countries with urban population between 0.5 SD and 1.5 SD. Group N show a significantly higher rate of deaths of COVID-19 compared to Group Y.

**Figure S10.**
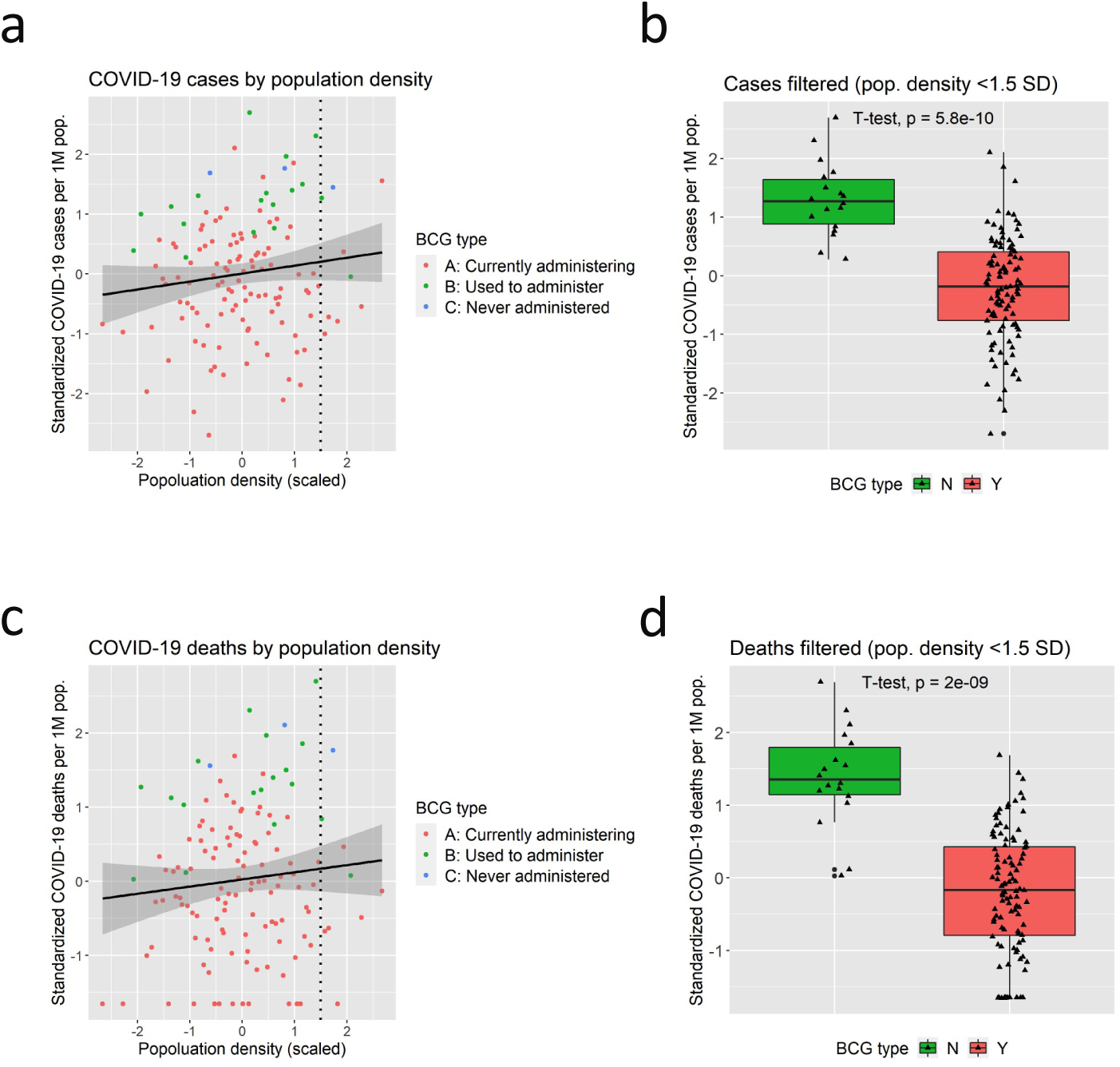
Correlations between population density and COVID-19: a. Scatter plot of total cases per one million population by population density (scaled). The vertical dotted line indicates the threshold of 1.5 SD, that is subsequently used for the analyses shown in the panel b. b. The boxplot of total cases per one million population sorted by BCG Group in countries with population density below 1.5 SD. Group N show a significantly higher rate of cases of COVID-19 compared to Group Y. c. Scatter plot of total deaths per one million population by population density. d. The boxplot of total deaths per one million population sorted by BCG Group in countries with population density below 1.5 SD. Group N show a significantly higher rate of deaths of COVID-19 compared to Group Y.

**Figure S11.**
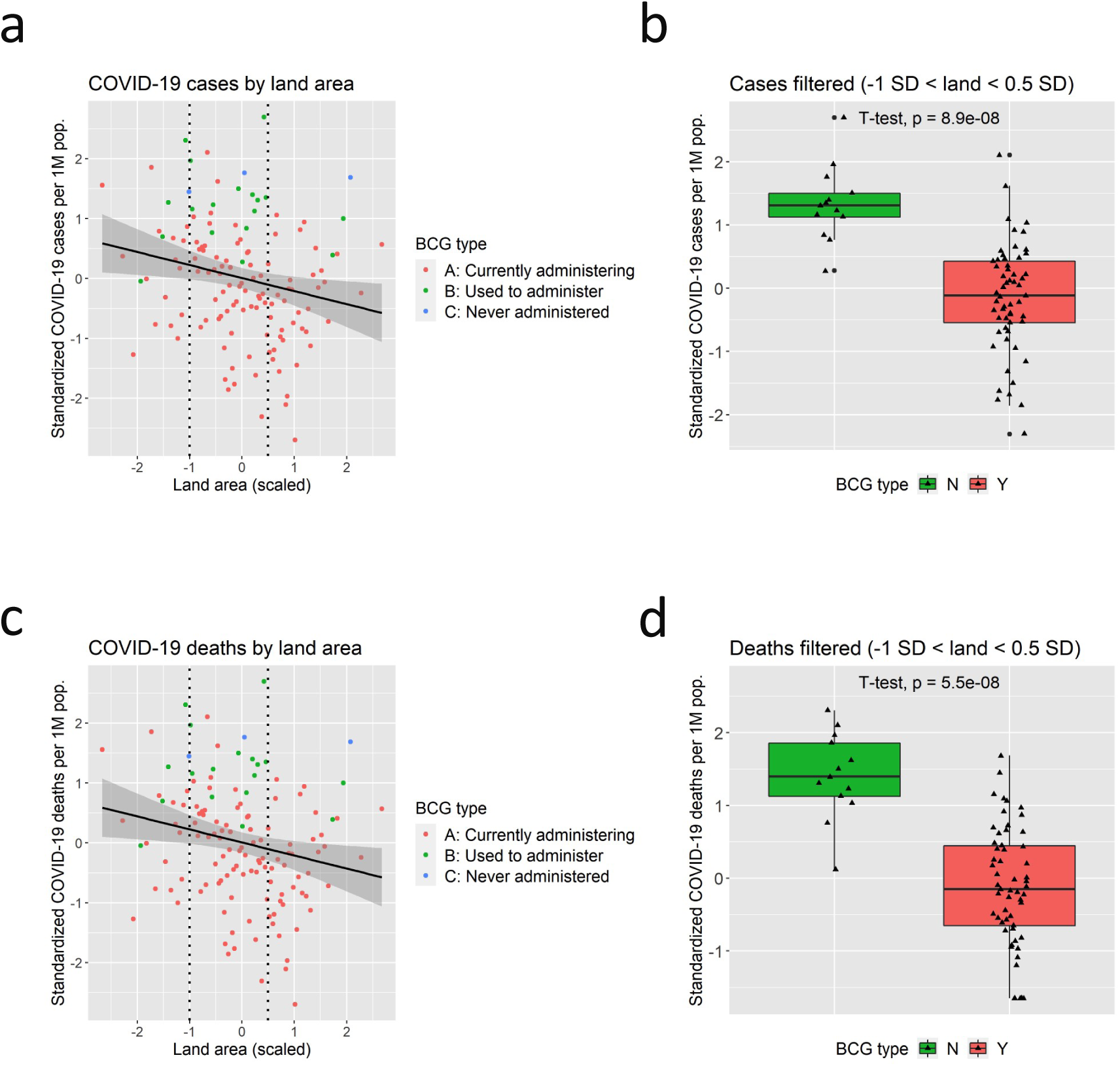
Correlations between land area and COVID-19: a. Scatter plot of total cases per one million population by land area (scaled). The vertical dotted line indicates the threshold of −1 SD and 0.5 SD, that is subsequently used for the analyses shown in the panel b. b. The boxplot of total cases per one million population sorted by BCG Group in countries with land area between −1 SD and 0.5 SD. Group N show a significantly higher rate of cases of COVID-19 compared to Group Y. c. Scatter plot of total deaths per one million population by land area. d. The boxplot of total deaths per one million population sorted by BCG Group in countries with land area between −1 SD and 0.5 SD. Group N show a significantly higher rate of deaths of COVID-19 compared to Group Y.

**Figure S12.**
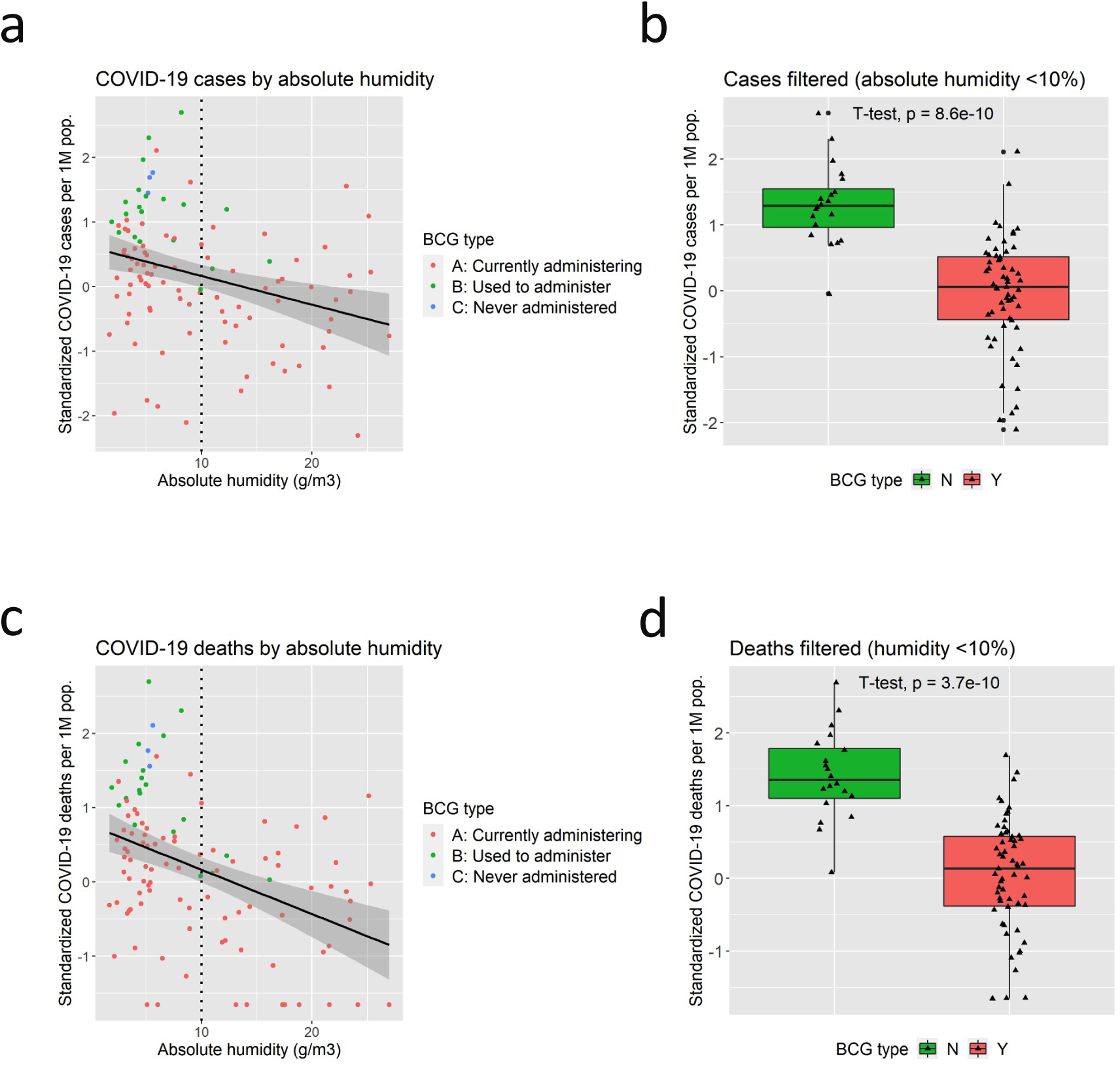
Correlations between absolute humidity in February and March and COVID-19: a. Scatter plot of total cases per one million population by absolute humidity (g/m^3^) in February and March. The vertical dotted line indicates the threshold of 10%, that is subsequently used for the analyses shown in the panel b. b. The boxplot of total cases per one million population sorted by BCG Group in countries with absolute humidity above 10 g/ m^3^. Group N show a significantly higher rate of cases of COVID-19 compared to Group Y. c. Scatter plot of total deaths per one million population by absolute humidity. d. The boxplot of total deaths per one million population sorted by BCG Group in countries with absolute humidity above 10 g/ m^3^. Group N show a significantly higher rate of deaths of COVID-19 compared to Group Y.

**Figure S13.**
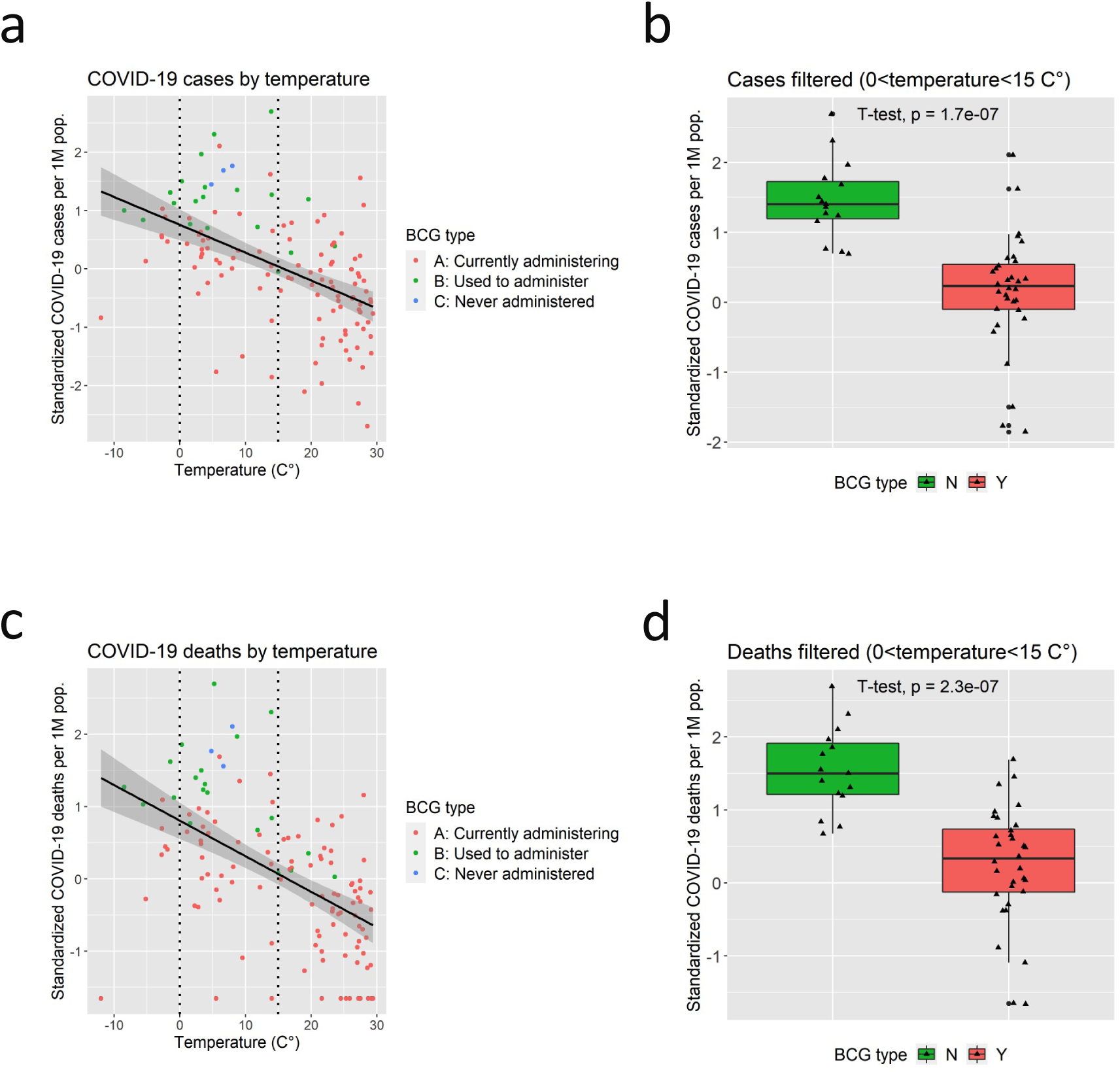
Correlations between temperature in February and March and COVID-19: a. Scatter plot of total cases per one million population by temperature (°C) in February and March. The vertical dotted line indicates the threshold of 0°C and 15°C that is subsequently used for the analyses shown in the panel b. b. The boxplot of total cases per one million population sorted by BCG Group in countries with temperature between 0°C and 15°C. Group N show a significantly higher rate of cases of COVID-19 compared to Group Y. c. Scatter plot of total deaths per one million population by temperature. d. The boxplot of total deaths per one million population sorted by BCG Group in countries with temperature between 0°C and 15°C. Group N show a significantly higher rate of deaths of COVID-19 compared to Group Y.

**Figure S14.**
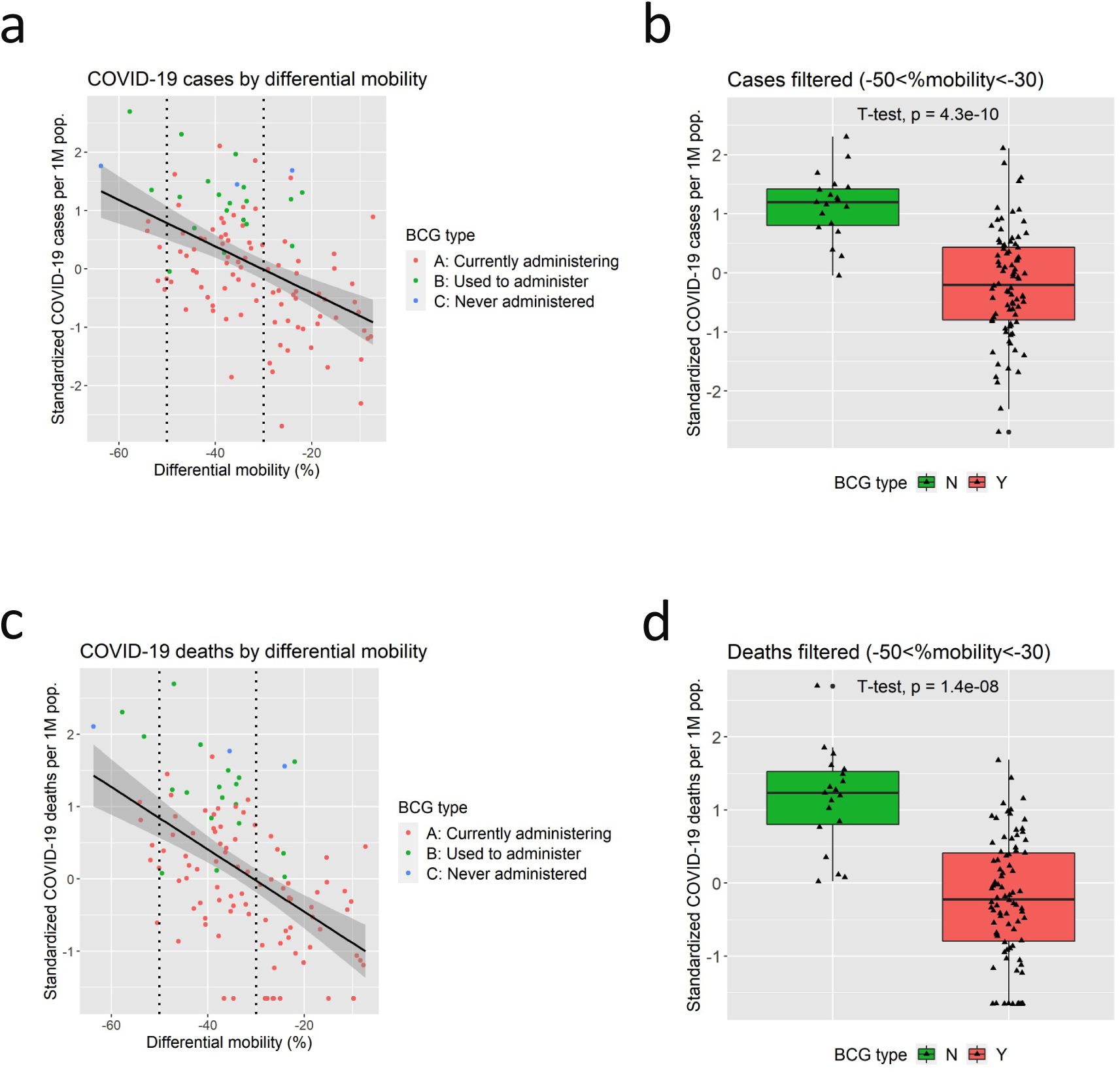
Correlations between community mobility change and COVID-19: a. Scatter plot of total cases per one million population by community mobility change from baseline based on COVID-19 community mobility reports by Google. The vertical dotted line indicates the threshold of −50% and −30% that is subsequently used for the analyses shown in the panel b. b. The boxplot of total cases per one million population sorted by BCG Group in countries with mobility change between −50% and −30%. Group N show a significantly higher rate of cases of COVID-19 compared to Group Y. c. Scatter plot of total deaths per one million population by mobility change. d. The boxplot of total deaths per one million population sorted by BCG Group in countries with mobility change between −50% and −30%. Group N show a significantly higher rate of deaths of COVID-19 compared to Group Y.

**Figure S15.**
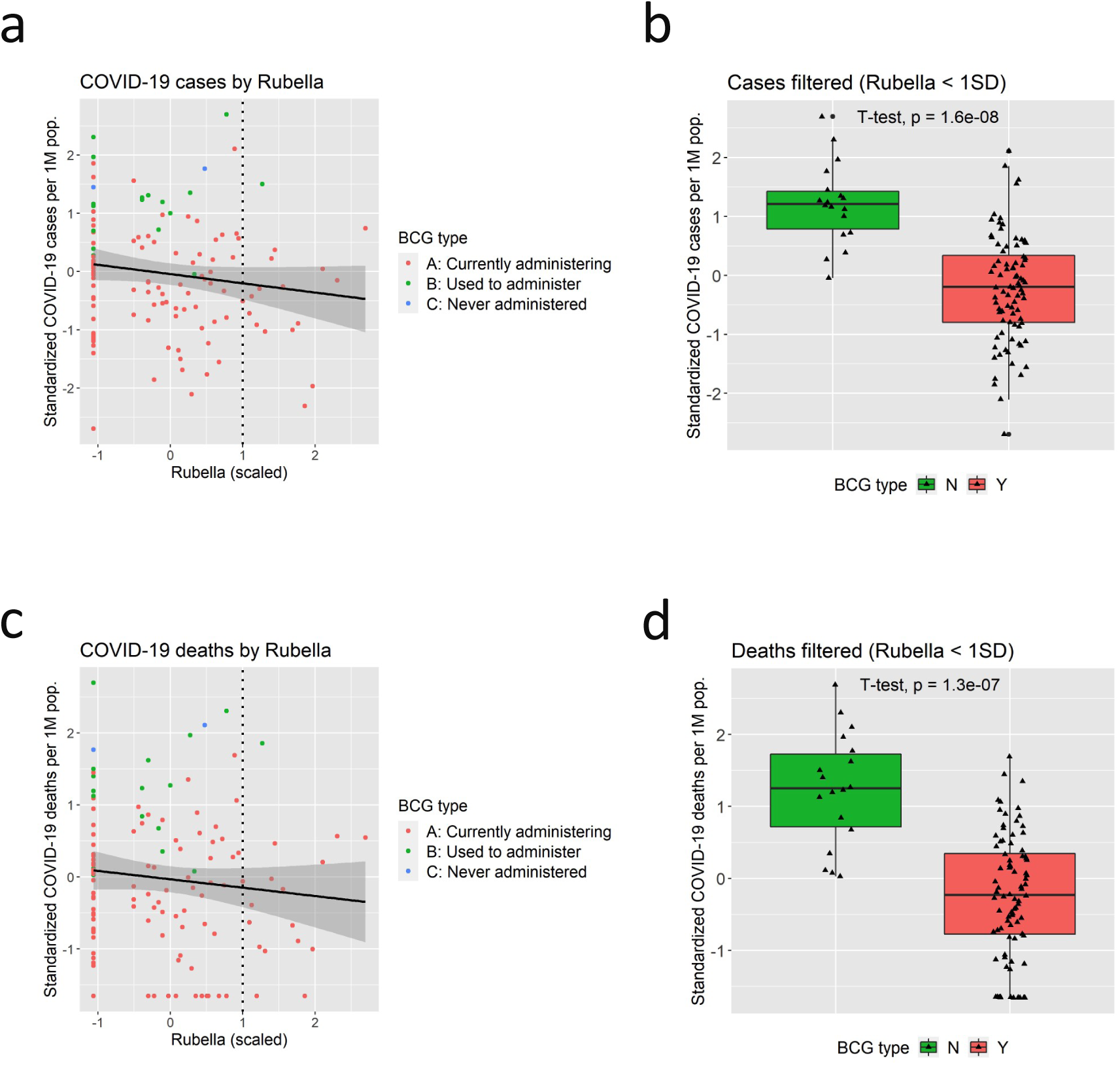
Correlations between incidence of Rubella and COVID-19: a. Scatter plot of total cases per one million population by incidence of Rubella (scaled). The vertical dotted line indicates the threshold of 1.0 SD, that is subsequently used for the analyses shown in the panel b. b. The boxplot of total cases per one million population sorted by BCG Group in countries with incidence of Rubella below 1.0 SD. Group N show a significantly higher rate of cases of COVID-19 compared to Group Y. c. Scatter plot of total deaths per one million population by incidence of Rubella. d. The boxplot of total deaths per one million population sorted by BCG Group in countries with incidence of Rubella below 1.0 SD. Group N show a significantly higher rate of deaths of COVID-19 compared to Group Y.

**Figure S16.**
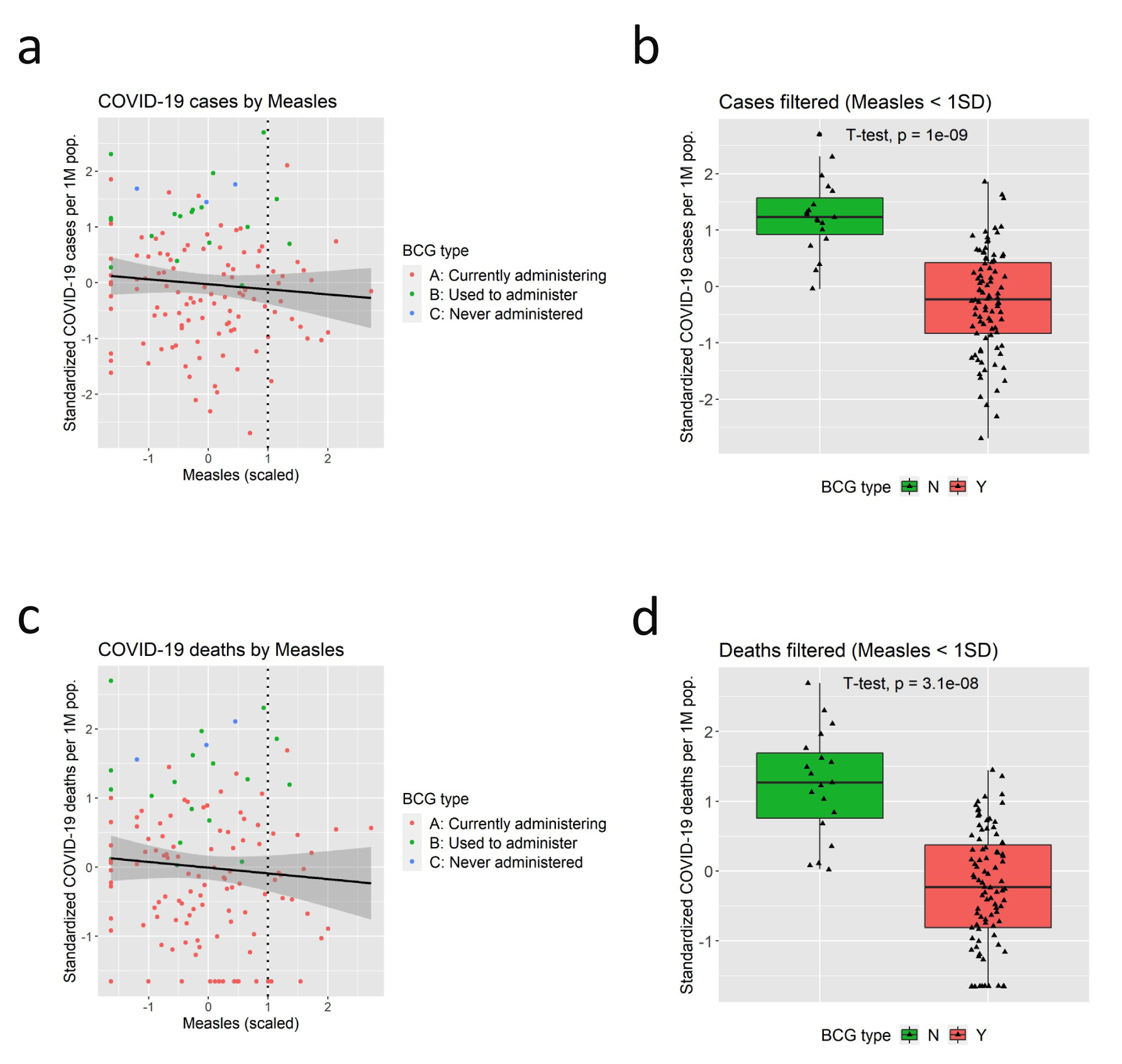
Correlations between incidence of Measles and COVID-19: a. Scatter plot of total cases per one million population by incidence of Measles (scaled). The vertical dotted line indicates the threshold of 1.0 SD, that is subsequently used for the analyses shown in the panel b. b. The boxplot of total cases per one million population sorted by BCG Group in countries with incidence of Measles below 1.0 SD. Group N show a significantly higher rate of cases of COVID-19 compared to Group Y. c. Scatter plot of total deaths per one million population by incidence of Measles. d. The boxplot of total deaths per one million population sorted by BCG Group in countries with incidence of Measles below 1.0 SD. Group N show a significantly higher rate of deaths of COVID-19 compared to Group Y.

**Figure S17.**
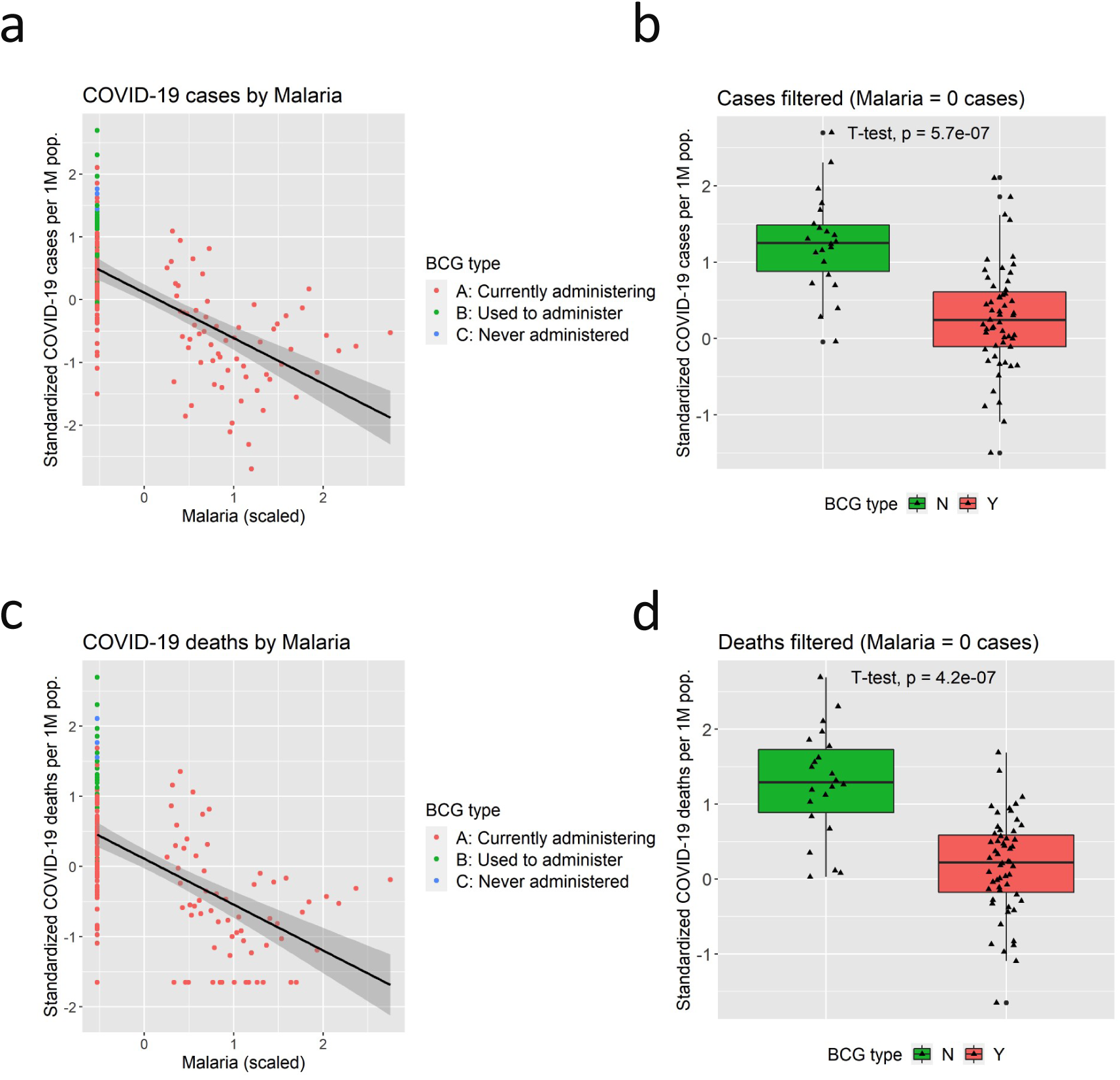
Correlations between incidence of Malaria and COVID-19: a. Scatter plot of total cases per one million population by incidence of Malaria (scaled). Note that there are countries that recorded no incidents of Malaria and such countries are displayed at extreme left side of the graph (a and c). b. The boxplot of total cases per one million population sorted by BCG Group in countries with zero incidence of Malaria. Group N show a significantly higher rate of cases of COVID-19 compared to Group Y. c. Scatter plot of total deaths per one million population by incidence of Malaria. d. The boxplot of total deaths per one million population sorted by BCG Group in countries with zero incidence of Malaria. Group N show a significantly higher rate of deaths of COVID-19 compared to Group Y.

**Figure S18.**
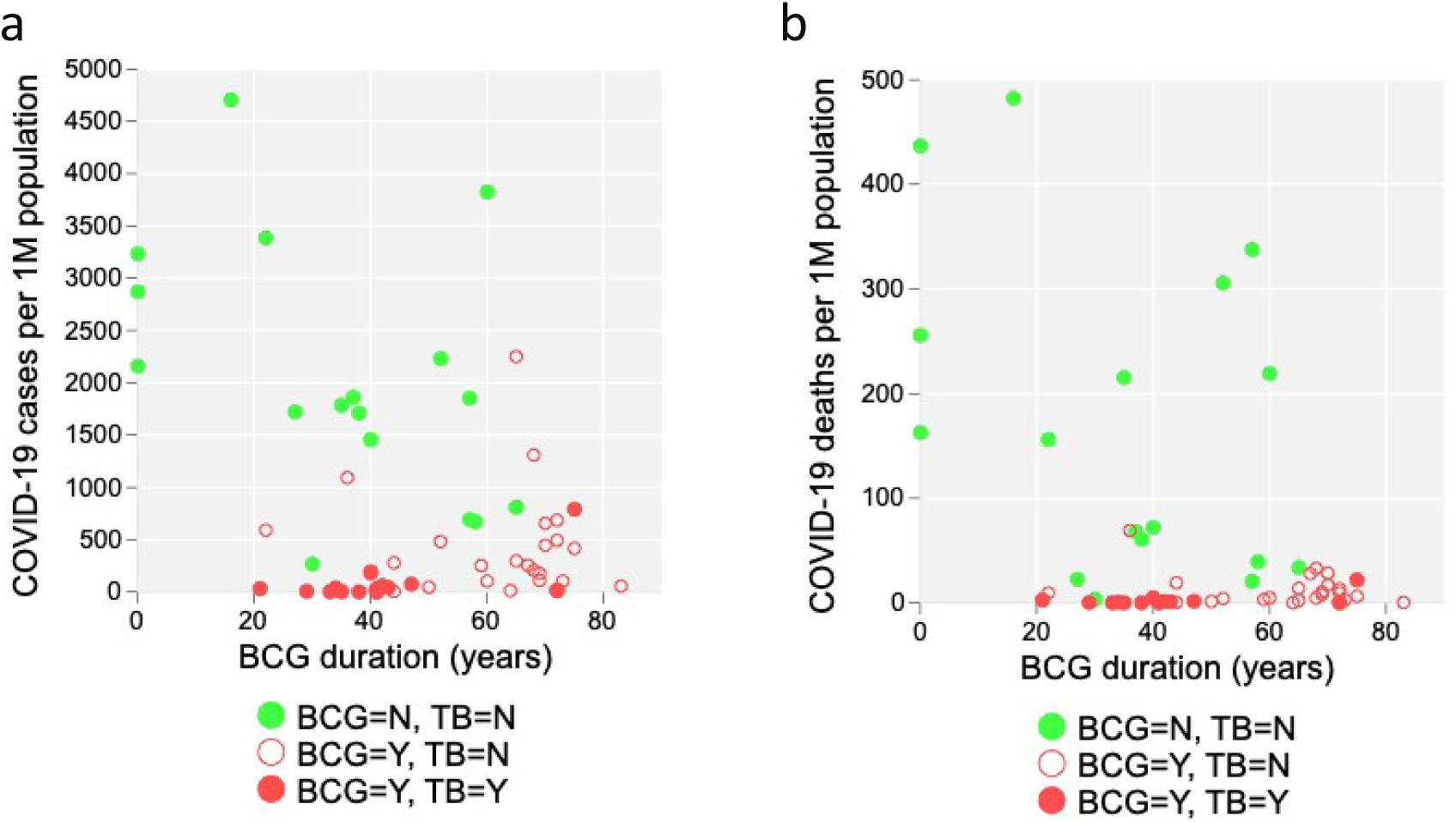
Duration of BCG (years) and COVID-19 cases/deaths: Scatter plot of total cases (a) or deaths (b) per one million population by BCG durations (years), during which each country had or has mandatory BCG vaccination policy. Green circles: Countries that discontinued mandatory BCG vaccination policy and do not suffer from high TB burden (less than 100 incidents per 100K population). Open red circles: Countries that have mandatory BCG vaccination policy and do not suffer from high TB burden. Closed red circles: Countries that have mandatory BCG vaccination policy and currently suffer from high TB burden. Note that only the countries whose duration of BCG could be obtained are shown in the graph.

**Figure S19.**
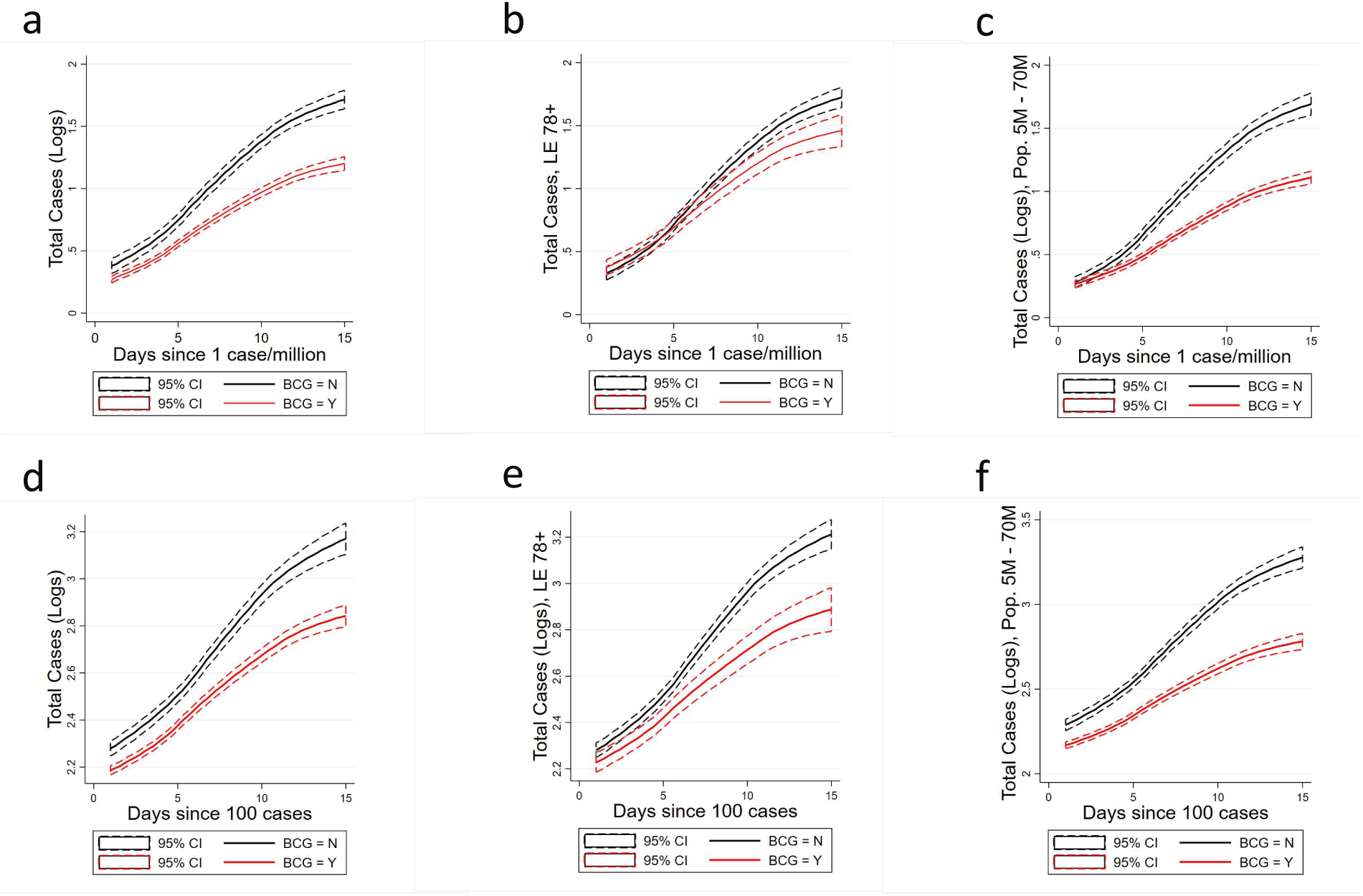
Growth curves, cases by subpopulations: mean log-transformed total cases by BCG Group. Panels a-c are for the first 15 days since a country reaches 1 case per million. Panels d-f are for the first 15 days since a country reaches 100 cases. Panels a & d present the full sample, Panels b & e restricts the sample to countries with life expectancy of 78 or higher, and c & f restrict the sample to countries with populations ranging from 5 Million to 70 Million. Group N exhibits a significantly higher growth rate of COVID-19 cases compared to Group Y.

**Figure S20.**
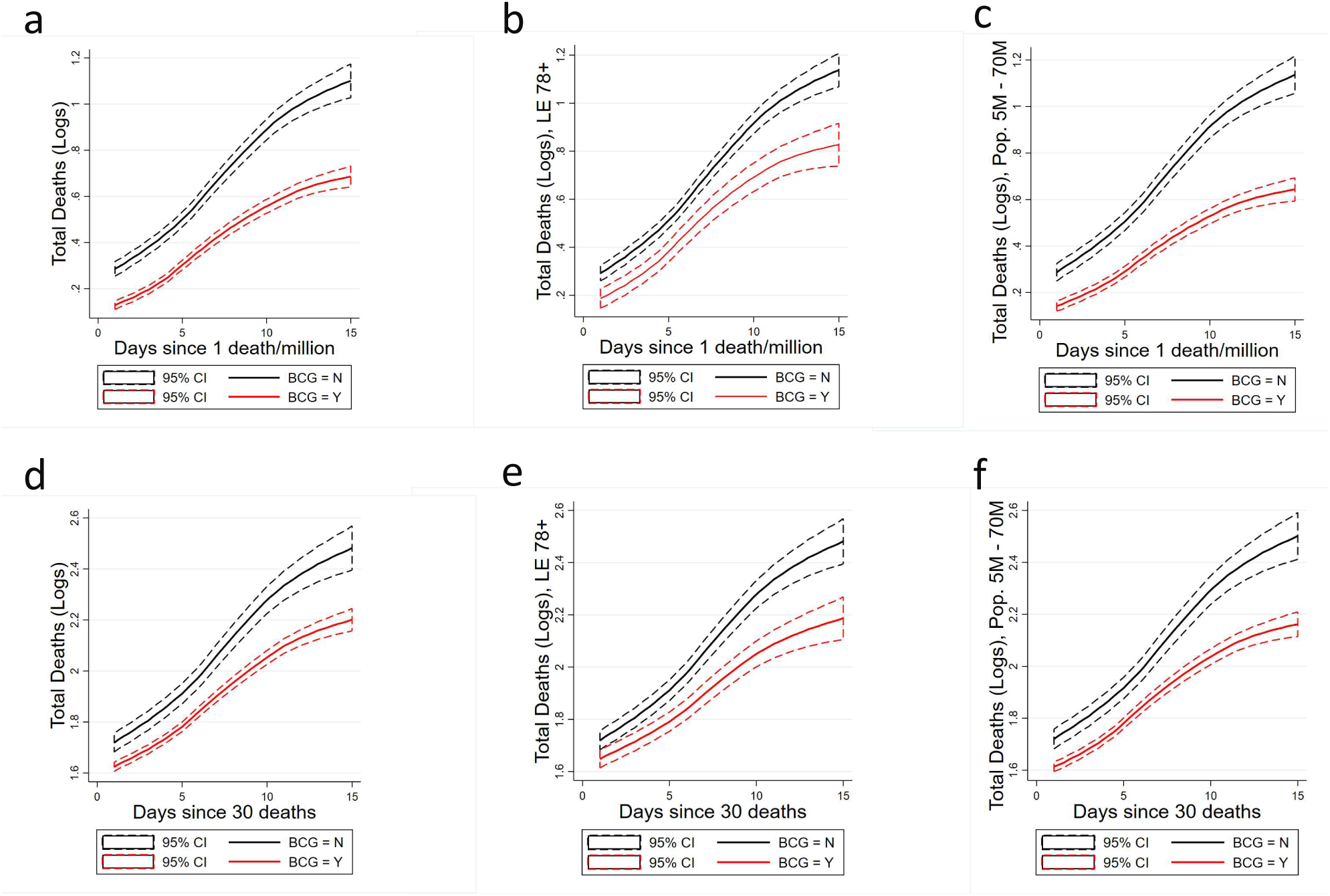
Growth curves, deaths by subpopulations: Mean log-transformed total deaths by BCG Group. Panels a-c are for the first 15 days since a country reaches 1 death per million. Panels d-f are for the first 15 days since a country reaches 30 deaths. Panels a & d present the full sample, Panels b & e restricts the sample to countries with life expectancy of 78 or higher, and c & f restrict the sample to countries with populations ranging from 5 Million to 70 Million. Group N exhibits a significantly higher growth rate of COVID-19 deaths compared to Group Y.

**Figure S21.**
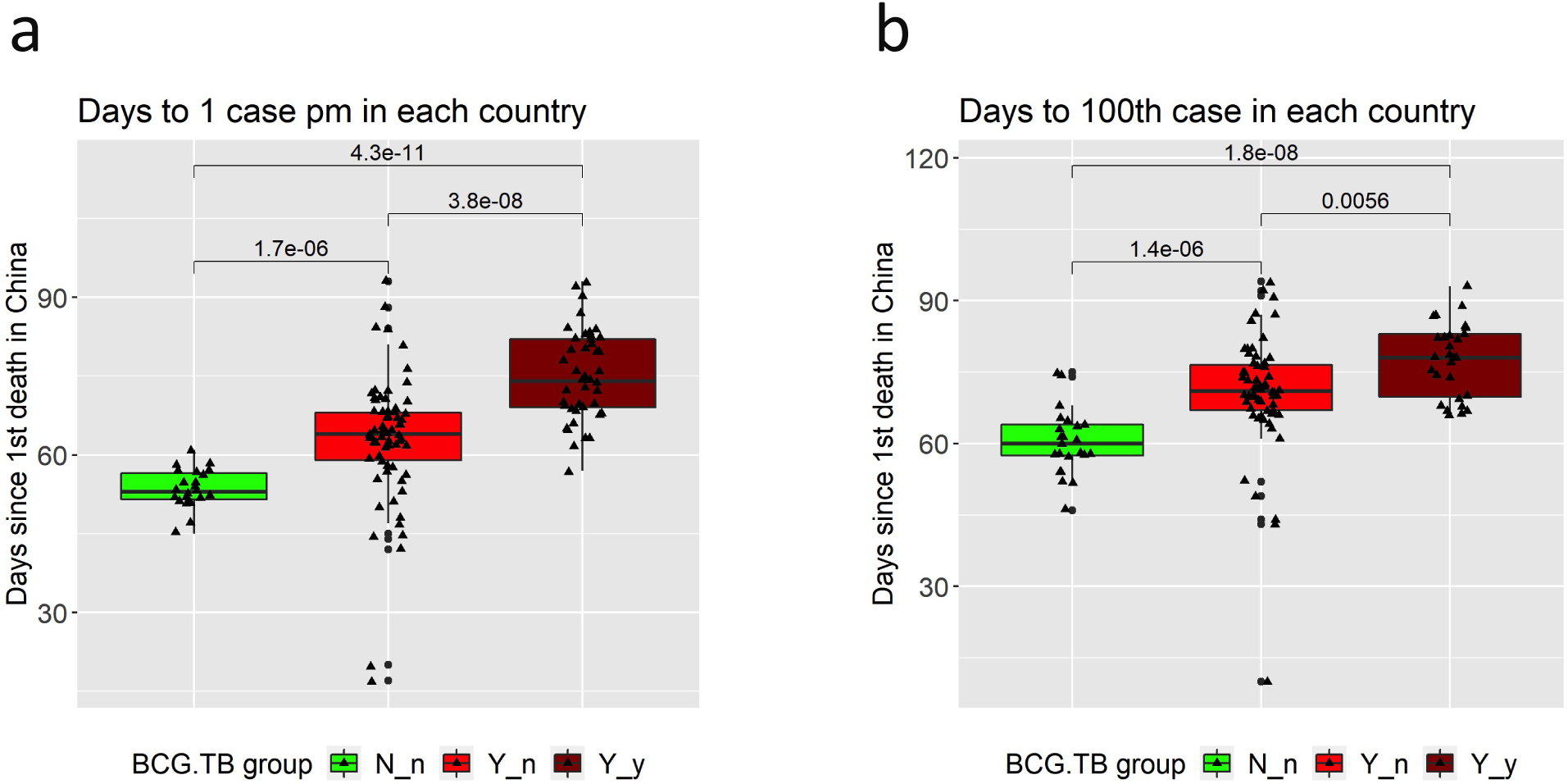
The number of days from 1^st^ death in China to 1 case per 1M population (a) or 100^th^ case (b) in each country was compared across 3 groups (Wilcoxon test; N_n group: The countries that do not have BCG vaccination policy at present and are not classified as high tuberculosis (TB) burden countries; Y_n group: The countries that have BCG vaccination policy at present and are not high TB burden countries; Y_y group: The countries that have BCG vaccination policy at present and are high TB burden countries). Highly significant differences between groups were found, suggesting that BCG policy and TB burden may have had effects on delaying the initiation of the propagation of infection.

